# Live wildlife trade in markets – a scoping review to inform risk assessment of emerging infectious diseases

**DOI:** 10.1101/2021.09.13.21263377

**Authors:** V.J. Brookes, O. Wismandanu, E. Sudarnika, J.A. Roby, L. Hayes, M.P. Ward, C. Basri, H. Wibawa, J. Davis, D. Indrawan, J. Manyweathers, W.S. Nugroho, S. Windria, M. Hernandez-Jover

**Author notes:** **Corresponding author** Victoria J. Brookes,. Sydney School of Veterinary Science, Faculty of Science, Camperdown, NSW, 2006, Australia.

## Abstract

Wet markets are important for food security in many regions worldwide but have come under scrutiny due to their potential role in the emergence of infectious diseases. The sale of live wildlife has been highlighted as a particular risk, and the World Health Organisation has called for the banning of live, wild-caught mammalian species in markets unless risk assessment and effective regulations are in place. Following PRISMA guidelines, we conducted a global scoping review of peer-reviewed information about the sale of live, terrestrial wildlife in markets that are likely to sell fresh food, and collated data about the characteristics of such markets, activities involving live wildlife, the species sold, their purpose, and animal, human, and environmental health risks that were identified. Of the 59 peer-reviewed records within scope, only 25% (n = 14) focussed on disease risks; the rest focused on the impact of wildlife sale on conservation. Although there were some global patterns (for example, the types of markets and purpose of sale of wildlife), there was wide diversity and huge epistemic uncertainty in all aspects associated with live, terrestrial wildlife sale in markets such that the feasibility of accurate assessment of the risk of emerging infectious disease associated with live wildlife trade in markets is limited. Given the value of both wet markets and wildlife trade and the need to support food affordability and accessibility, conservation, public health, and the social and economic aspects of livelihoods of often vulnerable people, there are major information gaps that need to be addressed to develop evidence-based policy in this environment. This review identifies these gaps and provides a foundation from which information for risk assessments can be collected.

## 1. Introduction

The COVID-19 pandemic – together with the emergence of SARS, Ebola virus disease and avian influenza H5N1 – has focused global attention on the potential role played by animal markets in the emergence of novel pathogens in human populations. Following the epidemiologic association between early cases of COVID-19 and the Huanan seafood market in Wuhan, Hubei, China (Chen et al., 2020; Huang et al., 2020), the potential for spill-over of potentially pathogenic microbes from live wildlife traded at fresh food markets, or ‘wet markets,’ was again highlighted as a risk for emergence of infectious diseases. Reactions have ranged from calls to improve risk assessment at wet markets to banning the sale of live wildlife in markets (World Health Organisation, 2021; Xiao et al., 2021). Although food-borne disease risks due to inadequate sanitation and hygiene practices at wet markets are well-documented (World Health Organization, 2006), the risk of pathogen spillover associated with live wildlife sale in wet markets and the circumstances in which this could lead to emergence of a new infectious disease are not well understood (Naguib et al., 2021; Pruvot et al., 2019).

Wet markets are common in many parts of the world, selling fresh food such as meat, seafood, vegetables and fruits, and sometimes live animals – including wildlife – which might be slaughtered and butchered at the market (Nadimpalli and Pickering, 2020; Sigal, 2020; Zhong et al., 2020). Such markets are found in China, Southeast Asia, Africa and South America (The Law Library of Congress, 2020). Food markets, including wet markets, serve local communities as social and commercial centres, and are especially important in urban settings in low income regions where they are important for food security by providing easy access to affordable food (World Health Organization, 2006).

Most emerging infectious diseases of humans have an animal origin (Jones et al., 2008; Taylor et al., 2001). Non-specific drivers of disease emergence – such as climate change and ecosystem degradation – are well-described, but exact mechanisms at interfaces between wildlife and other species such as livestock and people are generally poorly defined or unknown (Magouras et al., 2020; Plowright et al., 2017; Woolhouse et al., 2005). The probability of emergence of a pathogen depends on interacting factors associated with microorganism type, the characteristics and circumstances of the reservoir and potential new hosts, as well as opportunities for spill-over between these hosts by direct or indirect routes. These factors could facilitate or impede evolutionary processes and events that need to occur to enable cross-species transmission, and pathogenicity and transmissibility between individuals in the new host population (Plowright et al., 2017).

Wet markets provide an interface between live wildlife, domestic animals and people, and have previously been associated with disease emergence events (Woo et al., 2006). An outbreak of highly-pathogenic avian influenza (H5N1) in people in Hong Kong in 1997 was in part disseminated via live-bird markets (Tam, 2002). Crowding and mixing of bird species and unhygienic slaughter in markets, as well as proximity of markets to residential areas, were considered to be contributing factors. Similarly, live-poultry markets were considered a risk factor during an outbreak of H7N9 in people in Zheijiang, China (He et al., 2015). In the outbreak of severe acute respiratory syndrome (SARS) which emerged in late 2002 in Guangzhou Province, China, markets were again suggested as a risk factor because early community cases were more likely to have lived within walking distance of wet markets than community cases later in the outbreak (Xu et al., 2004). Supporting this, in markets in Guangzhou, there was evidence of greater exposure to SARS-CoV among live animal traders (seroprevalence 13.0%, N = 508), especially palm civet traders (seroprevalence 72%, N = 22), compared to control groups (seroprevalence range 1.2—2.9%, N = 284) (*Yu* et al., 2003). SARS-CoV-like virus was also isolated from live Himalayan palm civets (*Paguma larvata*) at a market in Shenzhen municipality, China, and evidence of infection with SARS-CoV-like virus was also found in a raccoon dog (*Nyctereutes procyonoides*), a Chinese ferret-badger (*Melogale moschata*), and people working at the market (Guan et al., 2003). Bats are considered to be natural reservoirs of many *Coronaviridae* and SARS-CoV-2 is closely related to a bat coronavirus (Andersen et al., 2020; Zhou et al., 2020). Although a route between bats and people has not been identified, species that are sometimes sold live in wet markets – for example, Pholidota (pangolin), one of the most trafficked animals in the world (Aisher, 2016) – have been suggested as intermediate hosts of SARS-CoV-2, though evidence for the role of any specific intermediate species is incomplete and consequently various theories remain controversial (Lam et al., 2020; Zhang et al., 2020). Additionally, evidence of the location or time during which transmission to humans could have occurred is lacking.

Whether live wildlife traded at markets provide opportunity for infectious disease emergence and the circumstances in which this might occur is of critical importance to developing and implementing policy to prevent pandemics. Risk assessments should form the basis of such policy by informing evidenced-based regulatory and surveillance frameworks that mitigate identified risks associated with the sale of live wildlife in food markets; such tools are already being developed (Wikramanayake et al., 2021). Detailed understanding of the hazards associated with live wildlife sold in markets is needed, not only to conduct meaningful risk assessments, but also to understand the policy environment of live wildlife trade in markets so that risk mitigation strategies can be implemented whilst supporting the essential socio-economic roles of markets.

In this review, our objective was to collate and describe peer-reviewed evidence of live, terrestrial wildlife sold in wet markets, including associated factors such as market type and documented risks to animal, human and environmental health. We aimed to provide a baseline of current knowledge – and identify gaps in knowledge – to support risk assessment and policy development regarding disease emergence associated with the sale of live, terrestrial wildlife at markets.

## 2. Materials and Methods

### 2.1 Protocol

This scoping review was conducted according to the Preferred Reporting Items for Systematic reviews and Meta-Analyses extension for Scoping Reviews (PRISMA-ScR) guidelines (Tricco et al., 2018). An overview of this methodology is described at http://www.prisma-statement.org/Extensions/ScopingReviews, accessed 18 October 2020.

The review protocol included three levels: Level 1, screening on title and abstract; Level 2, screening on full record; Level 3, data extraction. The web-based review platform Sysrev^©^ (Insilica LLC, 2020; www.sysrev.com, accessed 18 October 2020) was used for Levels 1 and 2, and a spreadsheet in Excel^©^ (Microsoft 2020, www.microsoft.com, accessed 18 October 2020) was used for Level 3.

We use the term ‘record’ to describe any bibliographic citation captured in the searches. We use the term ‘study’ as an individual investigation of wildlife sale in a market. Whilst most records comprised only one study, if a record contained multiple relevant studies (for example, theses, books, conference proceedings and reports), each relevant study was evaluated separately within that record.

### 2.2 Eligibility criteria

Types of records that were eligible included theses, dissertations, and peer-reviewed literature for which the full text was available. Records were eligible if they were published in English, from 1980 onwards, and from any country, and contained primary research of interest about the sale of live terrestrial wildlife at markets likely to sell food.

Markets of interest were collections of stalls which were identified in the record as food, wet or fish markets, street markets (when the meaning was not in the context of covert sale), medicinal markets, wildlife markets or open fairs. Pet shops, restaurants, bars or cafés were not included.

Terrestrial wildlife of interest were animals from classes Amphibia, Reptilia, Aves and Mammalia (see Supplementary Material for lists of orders in each class) which were presented live for sale at the market, and were not feral, companion or livestock animals. Vermin that lived in and around the market were not within scope unless presented live for sale at the market. Meat or other products from animals that were slaughtered prior to presentation for sale at the market were also not within scope. Live wildlife could be sold at the market for any purpose; for example, pets, medicine and consumption, including wildlife that were slaughtered at the market for these purposes.

Records which described the chain of events around wildlife trade (for example, hunting and sale of bushmeat) but did not include information about the sale of live wildlife at specific markets were excluded.

### 2.3 Information sources and search strategy

The literature search was conducted in July 2020, using the following combination of search terms:

i. market* OR sale,
ii. AND: wildlife OR “wild life” OR “wild-life” OR “wild animal” OR “wild bird”,
iii. AND: food OR medicine,
iv. timeframe: 1980—2020, language: “English.”

Three electronic databases were searched: Web of Science, PubMed, and SCOPUS. Searches for theses and dissertations were conducted in ProQuest Thesis and Dissertations Global Database, and EBSCO Open Dissertations. A literature search was conducted via the Google Scholar search engine, in which the first 100 results were screened. In addition, a search was conducted in Google Scholar with the addition of the term ‘TRAFFIC bulletin’ to identify relevant records from this journal, which was not available through the electronic databases.

All records were exported into the citation manager software Endnote (https://endnote.com/, accessed 18 October 2020), and duplicates were removed. Records were then uploaded to Sysrev© for Level 1 agreement tests and screening.

### 2.4 Selection of relevant records – Levels 1 and 2

Agreement tests were conducted prior to screening at Levels 1 and 2, in which all 10 reviewers in Levels 1 and 2 screened the same records (50 and 30 records at Levels 1 and 2, respectively). Conflicting opinions about inclusion or exclusion of records were discussed to achieve consensus between reviewers and to refine and improve the clarity of the questions at each level (see Supplementary Material 1).

During Level 1 (screening on title and abstract), two reviewers assessed each record. To maximise the sensitivity of identification of relevant records, records progressed to Level 2 if either reviewer assessed that the record might be eligible. During Level 2 (screening on the full record), two reviewers initially assessed each record. Records were only included for charting in Level 3 if there was consensus that the record met the eligibility criteria between at least 2 reviewers. Consensus was achieved via discussion and if required, consultation with a third reviewer.

### 2.5 Data items and charting process – Level 3

A spreadsheet was used to standardise data charting in Level 3 (Supplementary Material 1). Data items that were extracted included the year of publication, the location and year of the study, the driving concern for the study (for example, conservation or disease investigation), and then further data within the overarching topics of interest: markets, animals, and risks.

Data that were extracted about markets included the type of market (for example, wildlife or food), and the physical structure, frequency, and types of people and activities at the markets. Hygiene practices and temporal changes (for example, increased sale of live animal species) were also recorded.

Data that were extracted about animals at the markets included the species of live animals sold (if listed as individual species), their source, the volume sold and their physical condition and housing. Classes and orders of all wildlife (including dead animals and animal products) sold at the market were also extracted, as well as the total volume of all wildlife and the purposes of wildlife sale. Any non-wildlife species sold at the market were recorded, as well as contacts that were documented between wildlife species and any other live animal species.

Data that were extracted about risks included specific pathogens and diseases, measures of their frequency and how they were identified, and any other risks associated with human and animal health.

Due to the longer form at Level 3, four additional reviewers were included, and agreement amongst all reviewers was initially assessed with three records (selected based on diversity of study type). Modifications were made to the form following reviewer feedback. The form was then tested by all reviewers with two more records to assess agreement. All conflicts were discussed. Once consensus was achieved for all five records and further modifications to the Level 3 form were made to improve clarity and increase agreement, the full Level 3 review was conducted. Conflicting opinions about extracted data were discussed between each record’s pair of reviewers and a third reviewer to determine a consensus prior to synthesis of the extracted data.

### 2.6 Verification of the search strategy

Verification of the search strategy was undertaken by screening the bibliography list of three records that were retained for data analysis in Level 3 (one for each decade of the search window). Records which were potentially relevant to the study were included in the review process using the same methods as records identified in the initial search.

### 2.7 Synthesis of extracted data

Topics of interest were tabulated according to region, and information about each topic was then summarized and presented as a narrative.

## 3. Results

### 3.1. Screening

Initial and verification searches identified 3721 records. Following removal of 512 duplicates, 3209 records remained for screening (Figure 1). During Level 1 screening, 2795 records were excluded, leaving 414 records for screening of full text (Level 2). The most common reason for exclusion of records in Level 1 was lack of information about wildlife in markets (for example, the study investigated wildlife in the context of hunting or fishing; 98.6 %, n = 2757). During Level 2 screening, the most common reason for exclusion was lack of mention of live wildlife (92 %, n = 329). Of 133 records screened in the verification strategy, 5 were eligible for inclusion. Overall, 56 records were included for data charting and synthesis in Level 3 (Supplementary Material 2).

**Figure 1:**
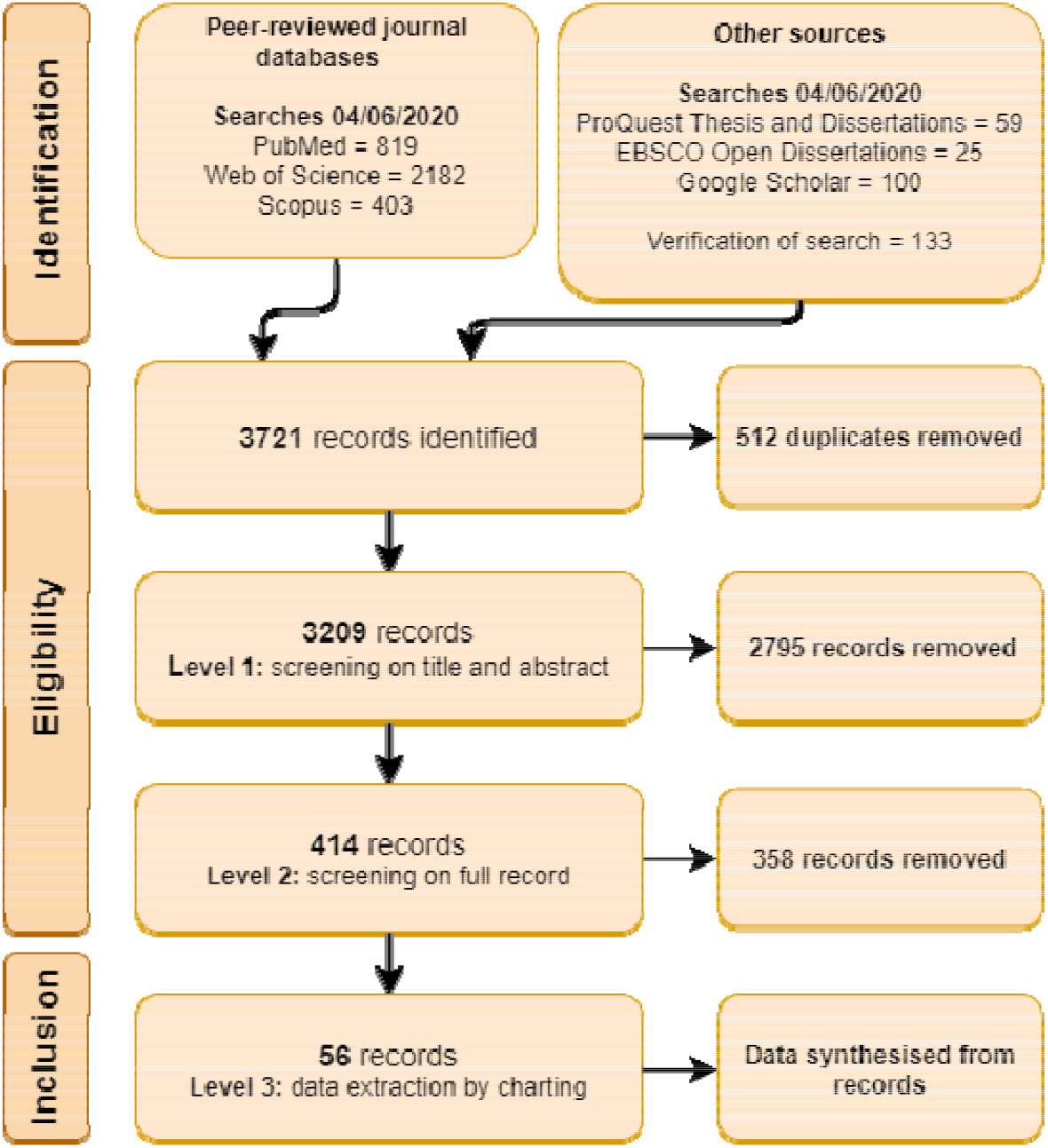
Diagram of the flow of records through the levels of a scoping review of reported research (1980–2020) associated with live, terrestrial, vertebrate wildlife sold for any purpose at markets likely to sell food.

### 3.2. Data charting

Of 56 records included in Level 3, all were single studies. A spreadsheet of the Level 3 studies and the data charted for each study is included in the Supporting Information. One record was sourced from a thesis (Edwards, 2012), and all other records were published in 35 peer-reviewed journals (Supplementary Material 3: Figure S1). The most frequently represented journal was *TRAFFIC Bulletin* (20%; n = 11), followed by *Biodiversity and Conservation* (7%; n = 4). Studies collected data from 28 countries (Figure 2). The most frequently represented study region was Southeast Asia (n = 22; mode country Lao PDR, n = 6), followed by China (here, classified as a region due to its large geographic size and number of studies; n = 17; mode province Guangdong, n = 7), Africa (n = 16; mode country Nigeria, n = 4), and South America (n = 8; mode country Brazil, n = 6). Two studies were conducted in more than one region: Europe and Africa (Spanish enclaves of Ceuta and Melilla, and Morocco; Nijman and Bergin (2017)), and China and Southeast Asia (Vietnam; Yiming and Dianmo (1998)).

**Figure 2:**
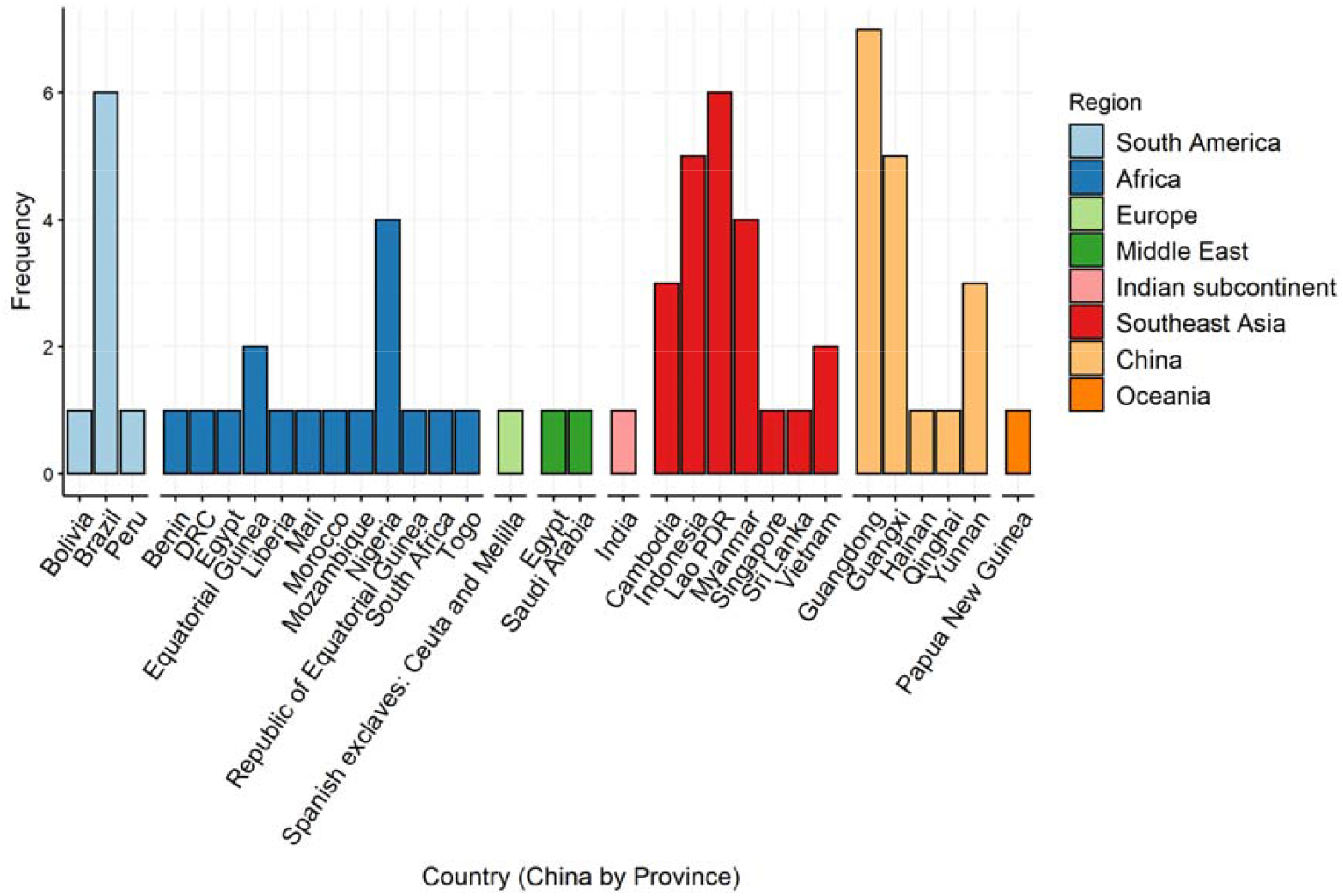
Regions and countries (China by Province) in which data were collected in peer-reviewed records identified in a scoping review of reported research (1980–2020) associated with live, terrestrial, vertebrate wildlife sold for any purpose at markets likely to sell food.

Studies used data collected over 1—21-year periods from 1980—2019 and were published from 1998—2020. Although publications increased after the avian influenza outbreak in Hong Kong in 1997, and then subsequent to the SARS outbreak in 2003, data in the publications in the early 2000s was frequently collected prior to 1997; therefore, research intensity did not appear to be influenced by particular EID events (Figure 3).

**Figure 3:**
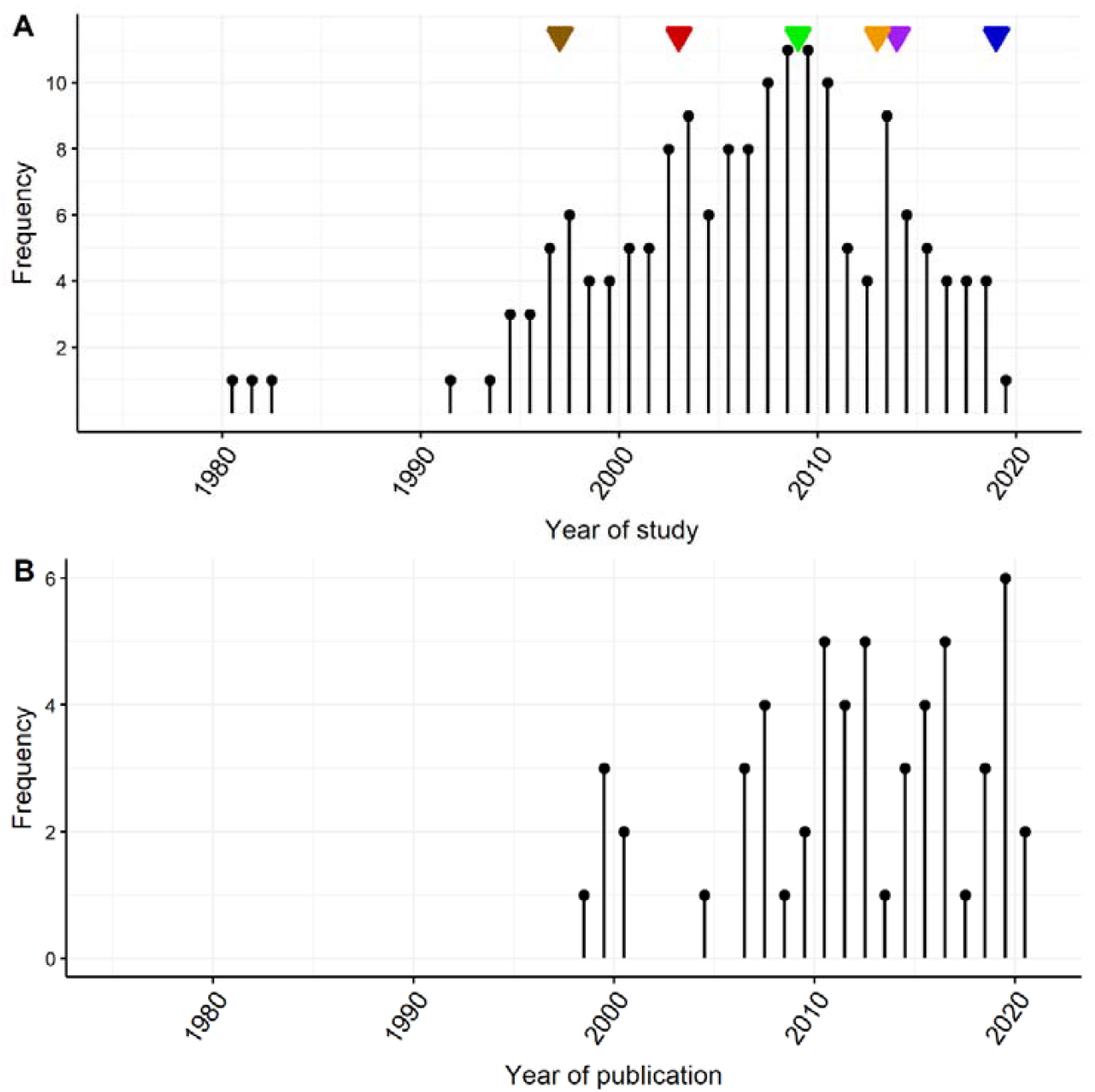
Years in which data were collected (A) and published (B) in peer-reviewed records identified in a scoping review of research associated with live, terrestrial, vertebrate wildlife sold for any purpose at markets likely to sell food. Triangles denote major outbreaks of emerging infectious disease: brown = Avian influenza A H5N1, Hong Kong 1997; red = SARS, 2003; green = H1N1, 2009; yellow = Avian influenza A H7N9, China 2013; purple = Ebola virus disease, West Africa 2007; blue = COVID-19, 2019.

The most common focus for studies was investigation of the wildlife trade in relation to conservation concerns such as species depletion and loss of biodiversity (75%; n = 42). Disease risk was the primary focus of study in only 14 studies (25%), and there was no association between the focus of studies and global region (X^2^ = 9.5, d.f. = 8, P = 0.3).

#### 3.2.1 Markets

The most frequent market types (according to author definitions and descriptions) were general markets in South America, traditional medicine and bushmeat markets in Africa, food in Southeast Asia and China, live-bird markets in Africa, and wildlife in Southeast Asia (Figure 4, and Supplementary Material 2). The frequency of topics associated with markets are shown in Table 1.

**Figure 4:**
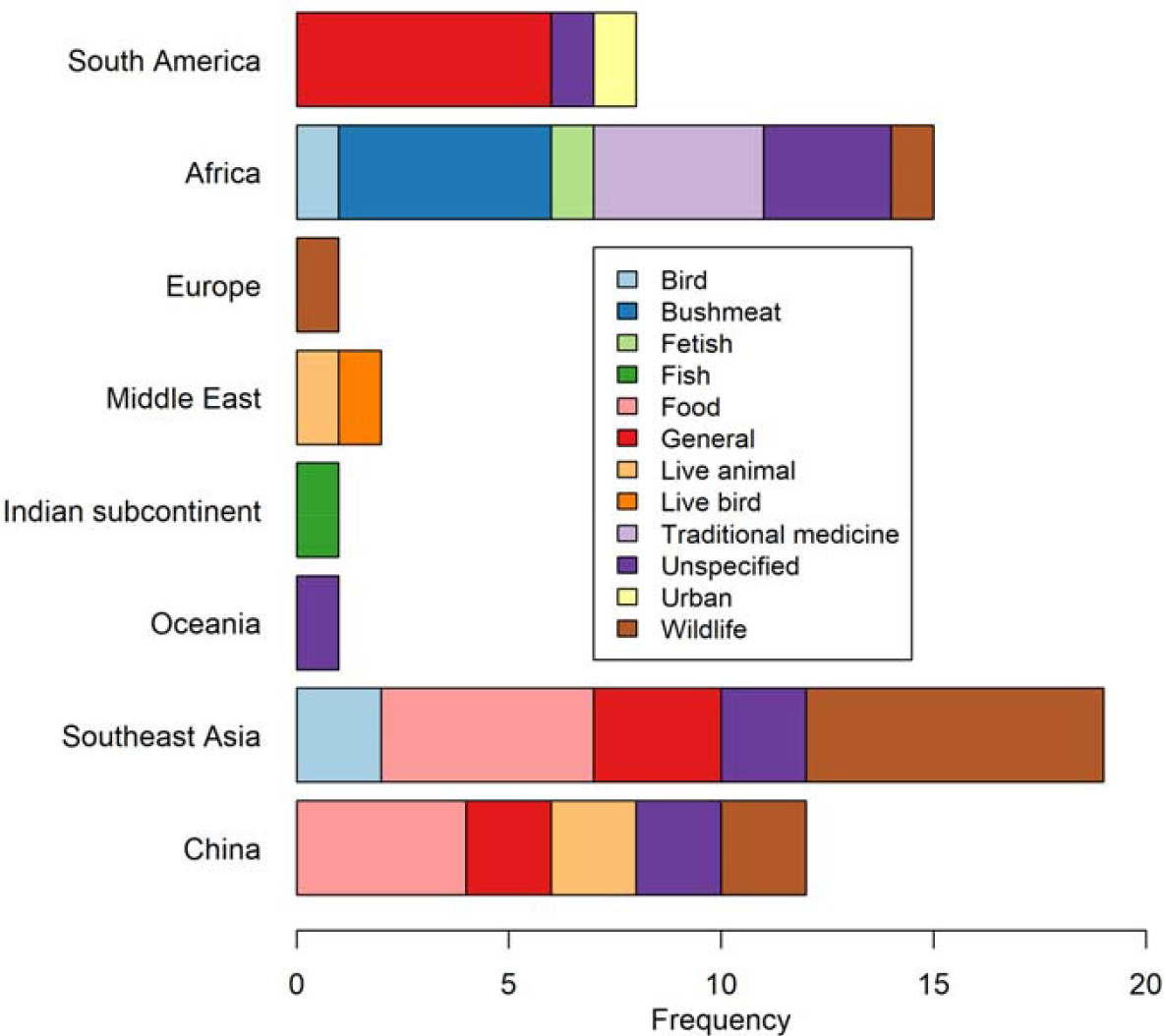
Market types identified in a scoping review of reported research (1980–2020) associated with the sale of live, terrestrial, vertebrate wildlife at markets likely to sell food globally.

**Table 1:**
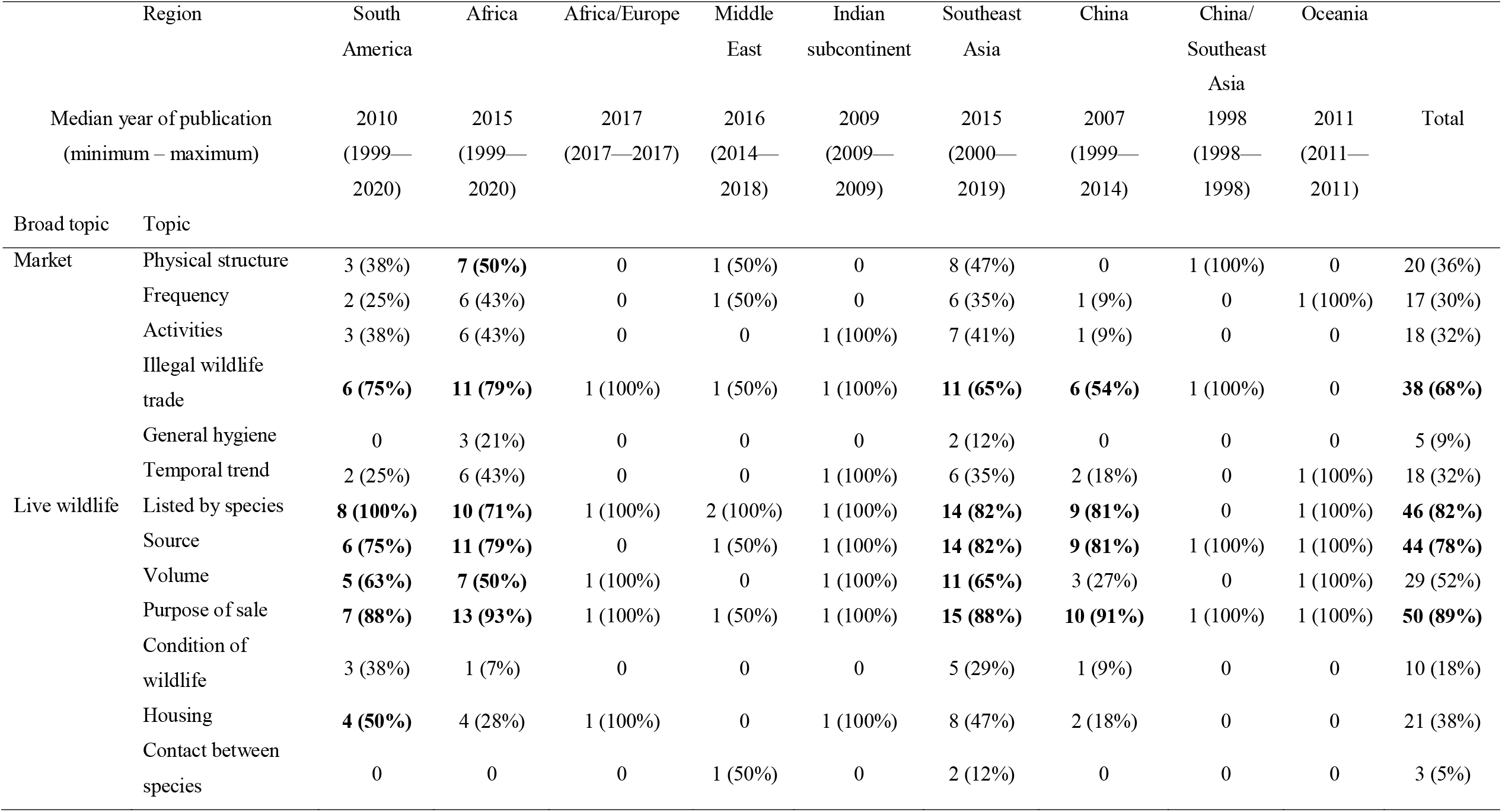
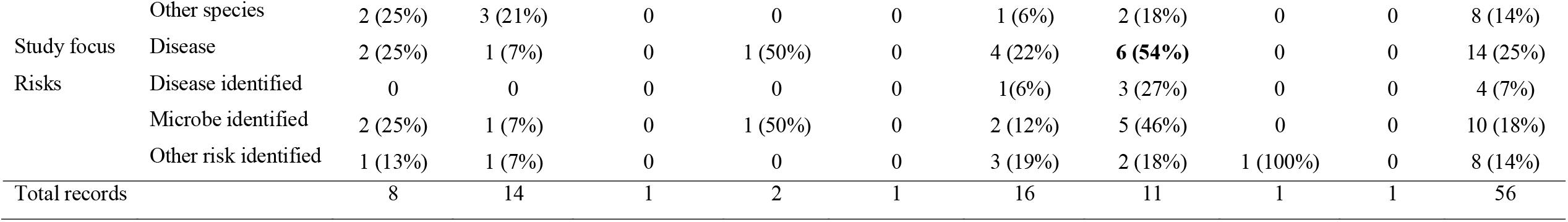
Number of records (percentage) from each region, which contained information from specific topics of interest in a scoping review of reported research (1980–2020) associated with live, terrestrial, vertebrate wildlife sold for any purpose at markets likely to sell food. Bold indicates that ≥ 50 % of studies from that region contained information on this topic (excluding regions in which there were ≤ 2 studies).

Market physical structure was described in 20 records (36%); a range of permanent and temporary structures were reported, either indoor or outdoor (for example, on the sides of roads or in car parks), and often in locations accessible for the arrival and distribution of goods. For example, in Belém, Brazil, market trade takes advantage of riverside ports (da Nobrega Alves and Pereira Filho, 2007), and in the Niger Delta, hunters deliver goods via rivercraft to one of the largest bushmeat markets in the region which is also accessible via recently constructed roads (Akani et al., 2015). In central Lao PDR, authors reported that the most active wildlife markets were located close to major roads which facilitated trade across Southeast Asia (Schweikhard et al., 2019).

Market frequency was highly variable although usually regular, with markets occurring every day on weekdays, on one day of the week only, on mornings only, at weekends only, at times dependent on the lunar calendar, or on special occasions. Unpredictable times and locations were a feature of some markets; for example, in Yunnan Province, China, wildlife trade could occur at irregular times and sites, and sometimes, markets appeared to be the sites of advertising and negotiation as vendors displayed their goods (including live wildlife in cages) whilst transfer of goods occurred elsewhere (Haibin and Kunming, 1999).

The types of people involved in markets were vendors and consumers – also called traders and buyers – as well as hunters and trappers, middlemen (also termed intermediaries and brokers), wildlife farmers, restaurant owners, government officials, slaughtermen, traditional healers and drivers. Middlemen could be involved in the chain of events before or after the market between wildlife acquisition and the end-user. For example, some middlemen traded between hunters and market vendors (these people could also be termed ‘collectors’ because they house wildlife as well as facilitated transactions), and others traded between market vendors and end-users such as restaurants, traditional medicine shops and tourists. Market trade was also associated with families as younger generations inherited the family business (for example, turtle collectors in Kerala, India; Krishnakumar et al. (2009)).

Illegal trade was reported in most records (68 %, n = 38), particularly if the focus was wildlife trade rather than disease risk (OR 17.0, 95 % CI 3.4—120.0, P < 0.001). Reports of illegal trade were not associated with a particular region (X^2^ = 5.1, d.f. = 7, P = 0.64). Types of illegal trade included the sale of wildlife species that were prohibited for sale by national or international laws, sourced from protected areas such as National Parks (for example, in Liberia and the Democratic Republic of the Congo and Liberia, reported by Greengrass (2016) and Van Vliet et al. (2012), respectively), collected by illegal methods such as hunting (for example, pangolin in Myanmar; McEvoy et al. (2019)), and illegally imported (for example, wildlife from Vietnam sold in Yunnan Province, China; Zhang et al. (2008)). Sometimes, laws were open to misinterpretation because they varied according to animal purpose (for example, hunting could be allowed depending on the destination and purpose of the wildlife in Indonesia; Chng et al. (2018); Janssen and Gomez (2019)) or were inconsistent between regional and national levels (for example, in Brazil; Fernandes-Ferreira et al. (2012)). Illegal trade was often conducted openly because it was either unregulated or ineffectively regulated. Examples include open trade despite signage that trade is illegal in Lao (Schweikhard et al., 2019), and turtle collectors in Kerala, India who were aware that trade was illegal, but were not deterred by small fines from local police and forest officials (Krishnakumar et al., 2009). Government officials were reportedly sometimes corrupt and therefore, not necessarily promoting regulatory activities (Haibin and Kunming, 1999). Sometimes illegal trade was clandestine. In Lao PDR, Davenport and Heatwole (2013) suspected that as live wildlife on display were sold, they were replenished from bags that were kept out of sight to avoid attention from local authorities, and in Mozambique, live wildlife and their products were concealed unless trade was likely (Williams et al., 2016).

Other than trade, activities directly associated with live wildlife included slaughter and butchering. This was most frequently reported in the context of reptile and amphibian trade (for example, in Africa, South America and Southeast Asia (Catenazzi et al., 2010; da Nobrega Alves and Pereira Filho, 2007; Edwards, 2012; Gilbert et al., 2012; Kusrini and Alford, 2006; Pruvot et al., 2019)). Levels of hygiene and sanitation were reported in only 6 records (11%) and were not more associated with studies which investigated disease risk than studies of trade (X^2^ = 2.7, d.f. = 1, P = 0.10). Cronin et al. (2015) observed that a bushmeat market in Bioko Island, Equatorial Guinea, Africa was occasionally closed for cleaning, and Van Vliet et al. (2012) noted that lack of refrigeration might be a reason for the high proportion of smoked bushmeat for sale at a market in Kisingani, DRC, Africa (and not an indication of a large catchment area of wildlife as some have previously suggested). Others also noted lack of refrigeration of fresh meat in studies in Asia (Pruvot et al., 2019; Shepherd and Nijman, 2007). In an assessment of zoonotic disease risk in markets throughout Lao PDR, (Greatorex et al., 2016) noted that hygiene practices such as hand-washing were rare, sanitation was poor (butchers knives and tables were rarely cleaned; blood and entrails were on the floor) and wildlife contact with fresh food was high in most markets. In a study from Nigeria and Egypt in which biosecurity compliance was evaluated (Fasanmi et al., 2016), wild animal trade was predictive for detection of highly-pathogenic avian influenza virus H5N1, and although not specific to activities involving wild animals, hygiene and sanitation measures were poor at many of the markets.

Temporal trends were mentioned in 18 records (32%), with changes in activity and species availability observed in some markets. In Yunnan Province, China, Haibin and Kunming (1999) noted increased smaller markets as well as underground wildlife trade networks. This could have been driven by increased regulation, reduced availability of some species, and increased consumption associated with development of the local economy and transport networks. Seasonality was observed in some markets; trade increased in the wet season in Nigeria (Akani et al., 2015) and in winter in Guangdong, China (Dong et al., 2007), and season was associated with species availability in Lao PDR and Brazil (Baía et al., 2010; Davenport and Heatwole, 2013; Schweikhard et al., 2019). Trade increased overall in Bioko Island (Cronin et al., 2015; Fa et al., 2000), and decreased in Togo, Mozambique and Papua New Guinea (D’Cruze et al., 2020; Eisemberg et al., 2011; Williams et al., 2016),whilst in markets in Myanmar, increases and decreases of particular species were observed over time (Min, 2012). Actors at markets also changed over time; for example, trade with non-locals has increased at Mong La market in Myanmar (Shepherd and Nijman, 2007). Trade also sometimes increased around religious festivals; for example, in Belém, Brazil (Nobrega Alves and Rosa, 2010).

#### 3.2.2 Wildlife Species

A total of 534 individual species of live, vertebrate, terrestrial wildlife were reported in the records from all taxonomic classes of vertebrates (Supplementary Material 3: Table S1). Of these, the most frequent orders were Testudines (37%, n = 197), followed by Passeriformes (20%, n= 109) and Squamata (12%, n = 65; Supplementary Material 3: Figure S2). Of mammalian orders that have been considered high risk for EID events (Delahay et al., 2021; Wikramanayake et al., 2021), no live species of Chiroptera (bats) were reported. Live Viverridae (large Indian, lesser Indian and masked palm civets) were reported in 4 records from China (Chow et al., 2014; Dong et al., 2007; Haibin and Kunming, 1999; Yiming and Dianmo, 1998). Pholidota (pangolins) were reported in 10 records from Africa (n = 5), Southeast Asia (n = 4) and China (n = 2) (Nijman 2016, (Akani et al., 2015; Edwards, 2012; Haibin and Kunming, 1999; Ingram et al., 2019; McEvoy et al., 2019; Shepherd and Nijman, 2007; Sodeinde and Soewu, 1999; Soewu and Ayodele, 2009; Yiming and Dianmo, 1998). Primate species were observed in markets in Africa, China and Southeast Asia, including a live chimpanzee in a bushmeat market in Liberia (Greengrass, 2016). Mustelidae species were observed in markets in China (Dong et al., 2007), and squirrels (Sciuridae) were observed in Southeast Asia and China (Pruvot et al., 2019; Yiming and Dianmo, 1998).

Most species were reported in records from China (51%, n= 271) and Southeast Asia (24%, n= 129), and the most frequent classes sold in China and Southeast Asia were Reptilia and Aves, respectively (Figure 5). Relatively few live species were reported in records from South America (n =73), or from the Middle East, Africa, Europe, the Indian subcontinent and Oceania (35, 29, 6, 2, and 1 species, respectively). In South America and the Middle East, most species were also of class Aves, and in the remaining regions combined, Reptilia were most frequently reported. Some records reported live wildlife by a common name only or by their genus (Figure 5 inset; Supplementary Material 3: Table S2). Of these, wildlife of class Aves were most frequently reported.

**Figure 5:**
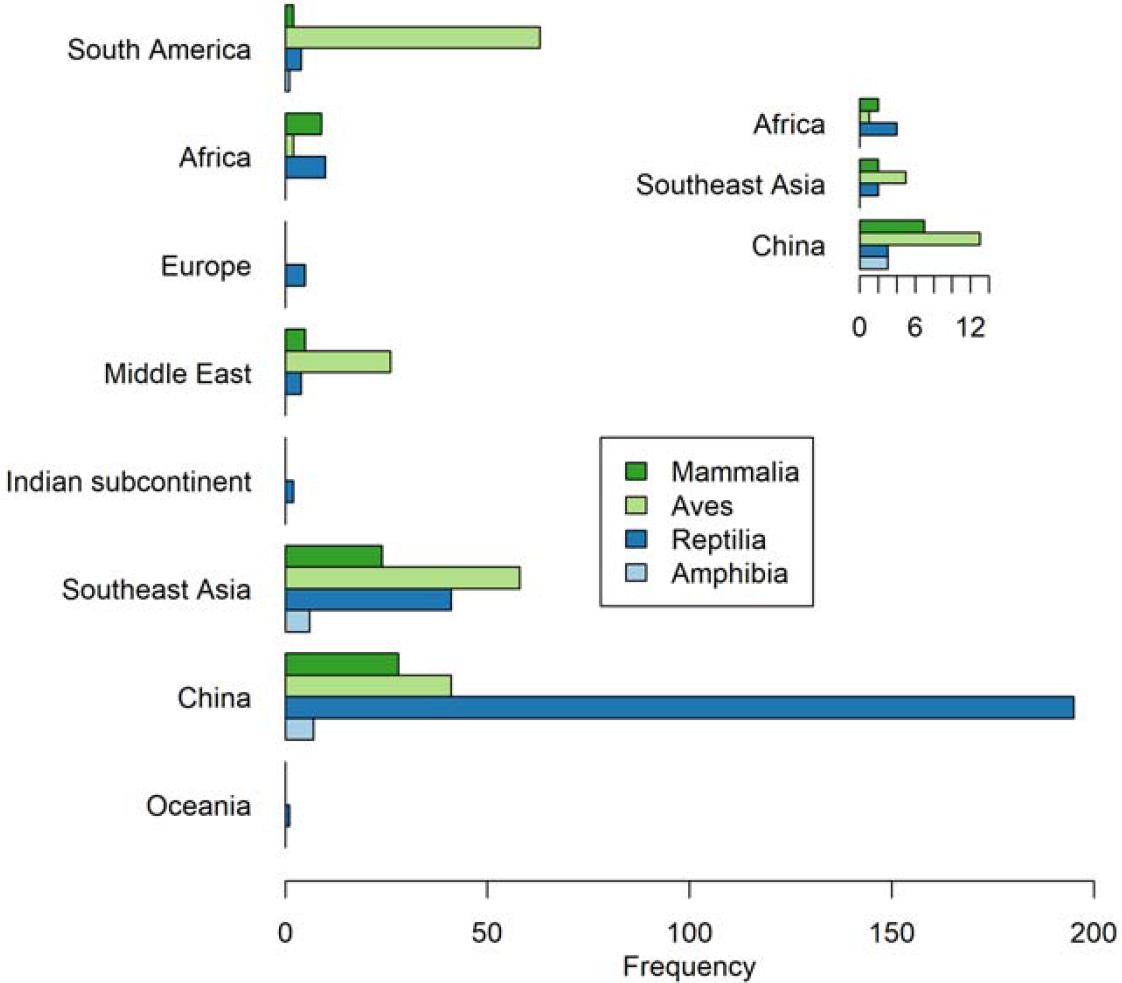
Classes of live, vertebrate, terrestrial wildlife reported in records in a scoping review of reported research (1980–2020) associated with the sale of live wildlife at markets likely to sell food globally. Main plot: classes of wildlife that were identified by species. Inset plot: classes of wildlife that were identified by common name, genus or order.

The greatest number of individual species of live, vertebrate, terrestrial wildlife reported in a single study was 157 turtle species reported from a 35-month survey of turtle trade in Hong Kong, Shenzhen and Guangzhou in southern China (Cheung and Dudgeon, 2006). Large numbers of live wildlife species were also reported from agricultural and general markets in Yunnan, China (n = 85; Haibin and Kunming (1999)), bird markets in Brazil (n = 51; Fernandes-Ferreira et al. (2012)), and wildlife markets in Sumatra (n= 48; Chng et al. (2018), at the Guangxi border between China and Vietnam (n = 47; Yiming and Dianmo (1998)), in South China (n = 26; Chow et al. (2014)), and in Tabuk, Saudi Arabia (n = 31; Aloufi and Eid (2014)). However, nearly half of the records only reported one live wildlife species (48%, n = 26; Figure S3).

Wildlife (live or products) were mostly sold for food and medicine globally, as well as pets in Southeast Asia and China, and spiritual purposes in South America and Africa (Figure 6). The reported volume of live wildlife and total wildlife products sold was dependent on the study design (number of markets and frequency of visits), and inventories were not possible in some markets due to covert trading – information from records about volume of wildlife is included in the Supplementary Material 2. Most studies reported that wildlife were sourced locally, often from national parks and similarly protected areas; however, in markets in Africa, the Middle East, China and Southeast Asia, wildlife (not necessarily live) were sometimes imported (Figure S4). The condition of live wildlife was rarely reported although Chow et al. (2014) noted tooth-like wounds of the feet of mammals that indicated they had been caught in traps, and Nekaris et al. (2010) found that the front teeth of lorises were clipped so that they made safer pets. In Lao PDR, Gilbert et al. (2012) observed animals tied with twine or bamboo through their hind limbs or foot webs, and Davenport and Heatwole (2013) observed frogs tied together, and lizards with their feet tied or their mouths wired or tied closed. In Brazil, Maloney et al. (2020) found that birds were in good condition. Live wildlife were often reported in unhygienic conditions in sacks, buckets in the case of reptiles and amphibians, or in overcrowded cages (Catenazzi et al., 2010; Nijman and Bergin, 2017; Stuart et al., 2000). The presence of other live animals was rarely reported (livestock in Mozambique; Williams et al. (2016), and poultry in Vietnam and China; Chow et al. (2014); Edmunds et al. (2011)). Likewise, proximity or contact between live wildlife and other species was rarely reported (contact with people; Pruvot et al. (2019), wild birds; Fasanmi et al. (2016), and poultry in the markets; Edmunds et al. (2011)).

**Figure 6:**
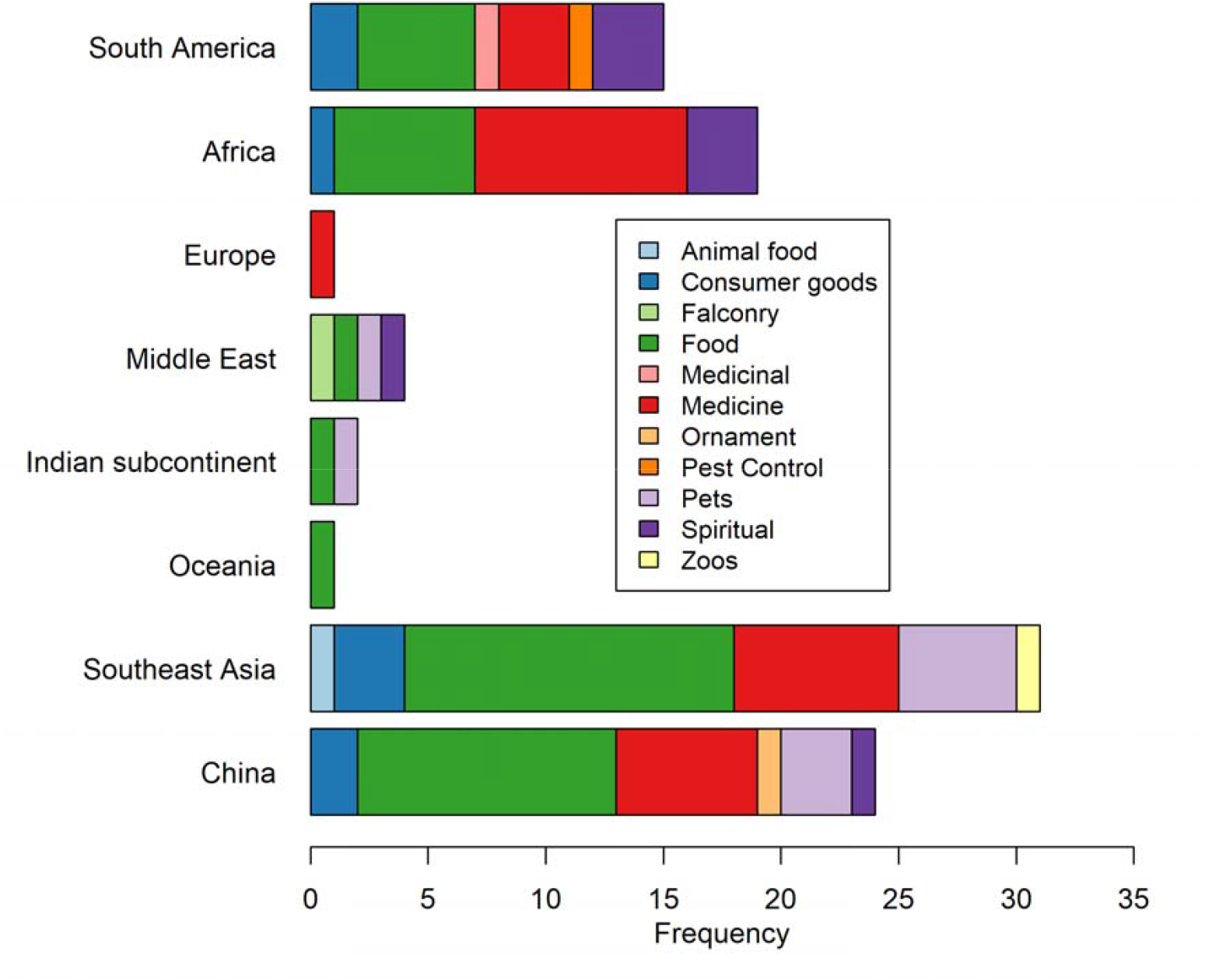
Purpose of sale of live wildlife and wildlife products reported in records in a scoping review of reported research (1980–2020) associated with the sale of live wildlife at markets likely to sell food globally. Main plot: classes of wildlife that were identified by species. Inset plot: classes of wildlife that were identified by common name, genus or order.

#### 3.2.2 Health risks associated with live wildlife

Twelve records identified specific diseases or microbes. Chytridiomycosis (caused by the fungus, *Batrachochytrium dendrobatidis*) was diagnosed by PCR and histological examination in 4 of 37 (10.8%) live bullfrogs in a food market in Yunnan Province, China (Bai et al., 2010). *B. dendrobatidis* was also detected in all live frogs sampled in a study in Peru (N = 5; Catenazzi et al. (2010)), and in one live bullfrog in Cambodia (N = 71; Gilbert et al. (2012)).

*Spirometra*, a zoonotic cestode that causes sparganosis, was detected with > 25% prevalence in snakes sampled in Guangdong (Wang et al., 2014; Wang et al., 2011) and Indonesia (Yudhana et al., 2019). In a study in markets in Brazil, the protozoan, *Blastocystis*, was detected with approximately 20% prevalence in Galliformes, Struthioniformes and Anseriformes, although not found in wild birds for sale (Maloney et al., 2020). Whilst some subtypes of *Blastocystis* have zoonotic potential, pathogenicity in people is variable; the authors noted that isolation of a subtype of *Blastocystis*, ST5, in ostriches in the study was significant because ST5 has been found in pigs and pig caretakers in China and Australia.

Eighteen isolates of avian influenza virus (H5N1, H9N2, H7N3, H7N9, H10N6, H3N6) were found in migratory Anseriformes in live-bird markets in Egypt (Kayed et al., 2019). Highly pathogenic H5N1 was also detected in tracheal and cloacal swabs, post-mortem, and moribund or freshly killed birds in 65% of live-bird markets surveyed in Nigeria and Egypt (N= 91; Fasanmi et al. (2016)).

In a study to examine the ecology and evolutionary pathways of *Coronaviridae* (CoV), Vijaykrishna et al. (2007) investigated viral sequences obtained from species including bats, humans and livestock, as well as wild animals (racoon dog and Chinese ferret badger) sampled at live-animal markets in Guangdong Province, China. The circumstances of the animals at the markets or the frequency of positive CoV identification was not described. In another study in live-animal markets in Guangdong and Guangxi Province, China, 24 live wildlife species were sampled (Dong et al., 2007). CoV was detected in several Asian leopard cats (*Prionailurus bengalensis*; n = 35/1,453, 2.4%) and Chinese ferret badgers (*Melogale moschata*; n = 11/934, 1.1%), and once in 6 other species: yellow-bellied weasel (*Mustela kathiah*), Siberian weasel (*M. sibirica*), masked palm civet (*Paguma larvata*), Chinese bamboo rat (*Rhizomys sinensis*), lesser Indian civet (*Viverricula indica*), and flying squirrel (*Petaurista sp*.). Ferret badgers and Asian leopard cats were available in the markets all year round, although most CoV positive swabs were detected in winter.

Zoonotic disease risks that were mentioned other than those associated with disease and pathogens detected in the studies included acknowledgement of the presence of large numbers of species susceptible to HPAI H5N1 in ornamental bird markets in Vietnam (Edmunds et al., 2011), and concern about zoonoses such as Ebola virus disease, rabies, and tuberculosis associated with handling primates in Benin, Africa (Sogbohossou et al., 2018). In markets in Guangxi at the China-Vietnam border, Yiming and Dianmo (1998) noted that wildlife mortality could be as high as 50%, especially during transport due to unhygienic conditions and overcrowding and that subsequent disposal of animals around trading areas and in forests could result in disease spread. Chow et al. (2014) commented that overcrowding could increase the probability of disease spread, and Kusrini and Alford (2006) noted that escaped exotic species (bullfrogs found in rice fields in West Java, Indonesia) could be a disease risk to local frog species.

## 4. Discussion

This review highlighted enormous global variability and epistemic uncertainty in all aspects regarding circumstances in which live, terrestrial wildlife are sold in markets that are likely to sell fresh food. Specific insights included the limited research focus on disease associated with live wildlife in markets, the wide variety of live wildlife species traded at markets especially in China and Southeast Asia, and the high accessibility of markets (both socially and geographically). These insights, as well as the variability and uncertainty related to the markets, the wildlife, and associated risks, greatly influence the feasibility of accurate assessment of the risk of emerging infectious disease associated with live wildlife trade in markets, and the development of effective, sustainable policy in this environment.

Most studies in this review focused on the impact of live wildlife trade in markets on conservation, particularly biodiversity loss. Whilst this can indirectly inform the scale of biodiversity loss as a broad driver of emerging infectious disease, focus on potential pathogens and disease is needed for risk analysis frameworks, which require information about specific hazards and exposure pathways. Many zoonotic and foodborne microorganisms have been identified in items other than live wildlife (including fresh meat, vegetables, and the environment) in wet markets worldwide, such as *Leptospira* spp., *Vibrio* spp., *Toxoplasma* spp., *Campylobacter* spp., and *Salmonella* spp. (Hamid et al., 2020; Kottawattage et al., 2017; Ngan et al., 2020; Sekoai et al., 2020), yet only three potentially zoonotic microorganisms (*Spirometra* species, *Coronaviridae*, and avian influenza virus) were reported from live wildlife in this review. There was also limited information about water, sanitation, and hygiene (WASH; https://www.who.int/health-topics/water-sanitation-and-hygiene-wash, accessed 20 August 2021) conditions associated with live wildlife trade, such as butchering and slaughter of wildlife, contamination of other food products or the environment, or contact between wildlife and other species, including people. Although market regulations are often aimed at reducing the risk of zoonotic and foodborne disease, and some countries – for example China, Indonesia, and Thailand – have specific WASH regulations applied to wet markets (The Law Library of Congress, 2020), there appears to be a gap in knowledge about the range of microorganisms and WASH risks associated with live wildlife. A greater understanding of current WASH conditions around activities associated with live wildlife trade (including market infrastructure such as hand washing facilities, adequate drainage, and separation of animals) is required to inform risk assessment exposure pathways. In addition, previous recommendations to increase surveillance of wildlife to identify potential pathogens and to detect and respond to emerging infectious disease (Grogan et al., 2014; Kuiken et al., 2005; Vrbova et al., 2010) appear to have been largely overlooked in the context of live wildlife in markets and should be adopted more comprehensively.

The greatest number of live wildlife species was reported from studies in markets in China and Southeast Asia. Wide species diversity might provide more opportunity for EID events due to a greater number of potential reservoir-spillover host pairs. However, following previous patterns of disease emergence, species from classes Mammalia and Aves have been considered a higher disease-risk due to their association with potentially pathogenic microorganisms (Olival et al., 2017; White and Razgour, 2020; Woolhouse et al., 2012), and a greater focus on these classes of wildlife might be warranted. Consistent with this, the World Health Organization have recommended suspension of trade of live-caught mammalian wildlife in traditional food markets (World Health Organisation, 2021). In this review, live mammalian wildlife species that could be considered high risk were identified in studies from markets in Africa, Southeast Asia, and China, and wild birds were also reported in markets from South America and the Middle East in addition to these regions. However, the likelihood of spillover will also depend on additional characteristics of the wildlife-potential spillover host interface, including its magnitude (amount of live wildlife traded and contact with the potential spillover species) and factors that affect transmission, such as stressors that might influence the shedding of microorganisms (including travel, overcrowded housing, comorbidities and the physical condition of the wildlife), the type of contact occurring and the susceptibility of the potential spillover host (Plowright et al., 2017; Wikramanayake et al., 2021). The scale of interfaces could not be determined in this review, and although most studies and species were identified from China and Southeast Asia, this might not be representative of the global distribution of markets or volume of live wildlife sold. Ultimately, resources for mitigation strategies will be limited, and development of effective policy will require assessment of relative risk dependent on the diverse global contexts that were apparent in this review, such as market type (for example, markets in this review were predominantly bushmeat in Africa, wildlife in Southeast Asia, and food in China) and species sold, as well as local, market-specific factors.

The scope of policy to mitigate EID risks associated with trade of live wildlife in markets is also important to define. If the goal is to prevent dissemination of disease from markets and an epidemic – and not only to prevent spillover of microorganisms from live wildlife reservoir hosts – social and geographic pathways from markets also need to be considered in risk assessments. Short value-chains that support local producers and ensure a supply of fresh food at low cost – due to the limited requirement for transport and refrigeration – have been considered an advantage of wet markets (Goldman et al., 1999; Zhang and Pan, 2013).

However, several studies in this review described how markets were located to facilitate trade and distribution, and highlighted that wildlife trade is a high-value, global activity with long value chains and movement of wildlife across many regions. In 2017, the value of the global wildlife trade was an estimated USD23 billion (van Uhm and Wong, 2019). They also described a multitude of people associated with live wildlife trade, consistent with the typologies defined by Phelps et al. (2016). Long value-chains which involve many people might be specific features of markets selling live wildlife that could be related to the purpose and value of wildlife. Although quantitative methods can be used to characterise risks and their pathways within markets (such as species abundance and contact durations), social science and ethnographic methods, such as those described by Edwards (2012) in the context of markets in Mali, Africa, are likely to be of greater value to understand complex wildlife trading networks. Understanding the drivers and risks behind network structure is valuable to identify potential control points (Phelps et al., 2016), and will also serve to understand the socio-economic impacts of regulations to mitigate risks. Networks involving people and transport are often small-world networks and it is possible that wildlife trade networks are of this type; such networks can be robust in the face of disruption (Hu and Verma, 2011). Therefore, as well as not addressing the drivers of wildlife consumption, banning trade of live mammalian wildlife in markets might not be an effective strategy to mitigate EID risks. It has already been suggested that those whose livelihoods depend on wildlife trade are likely to find alternative trade routes that avoid regulations; trade diverted to hidden networks could inadvertently pose a greater risk to public health (Roe et al., 2020).

Overall, this review demonstrates that accurate assessment of the risk of EIDs associated with live wildlife trade in markets is currently infeasible due to massive epistemic uncertainty. The magnitude of live wildlife-potential spillover host interfaces is unknown, high-risk species and microorganism pairs are largely unknown, there is limited information about activities that could influence exposure to microorganisms, and there is limited evidence to associate EID events with wet markets. Although this review did not identify records in languages other than English, the small proportion of records which focused on disease indicates that assessing EID risks in markets has had less research focus than the conservation impacts of live wildlife trade. This does not mean that markets are not a risk for EID emergence and that a precautionary approach is unwarranted. Ultimately however, policy will need to be evidence-based and commensurate with risk so that it can be implemented and sustained. The high-profile association of early COVID-19 cases with a wet market in Wuhan and the current recommendation to ban trade of live mammalian species in markets has catalysed the need to understand these risks. This review provides a baseline of currently available, peer-reviewed information about live wildlife trade in markets globally. Addressing the gaps in knowledge to accurately assess risks across the wide range of market types, activities and wildlife species purpose globally will provide the framework in which policy to promote biodiversity, protection of livelihoods and prevention of EIDs can be supported.

## Data Availability

All data are included in Supplementary Material

## Acknowledgments

We thank the Australian Department of Education, Skills and Employment, Enabling Growth and Innovation Program for their funding and support.

