## Supplementary Material 1 for "Live wildlife trade in markets – a scoping review to inform risk assessment of emerging infectious diseases"

V.J. Brookes^1, 2^, O. Wismandanu^3^, E. Sudarnika^4^, J.A. Roby^2, 5^, L. Hayes^1,2^, M.P. Ward^6^, C. Basri^4^, H. Wibawa^7^, J. Davis^8^, D. Indrawan^9^, J. Manyweathers^1, 2,^ W.S. Nugroho^10^, S. Windria^11^, M. Hernandez-Jover^1, 2^

^1^ School of Animal and Veterinary Sciences, Charles Sturt University, Australia

^2^ Graham Centre for Agricultural Innovation, (NSW Dept. of Primary Industries and Charles Sturt University), Australia

^3^ Veterinary Medicine Study Program, Faculty of Medicine, Padjadjaran University, Indonesia

^4^ Faculty of Veterinary Medicine, IPB (Institut Pertanian Bogor) University, Indonesia

^5^ School of Biomedical Sciences, Faculty of Science, Charles Sturt University, Australia

^6^ Sydney School of Veterinary Science, The University of Sydney, Australia

^7^ Disease Investigation Centre Wates, Directorate General of Livestock and Animal Health Services, Ministry of Agriculture of Indonesia

^8^ Australian Department of Agriculture, Water and the Environment, Canberra, Australia

^9^ School of Business, IPB University, Indonesia

^10^ Faculty of Veterinary Medicine, Universitas Gadjah Mada, Indonesia

^11^ Department of Biomedical Sciences, Division of Microbiology, Veterinary Medicine Study Program, Faculty of Medicine, Padjadjaran University, Indonesia

Forms used at each level of the scoping review

Level 1 Screening on title and abstract:

| Question | Inclusion value |
| --- | --- |
| Does the record include information about live terrestrial wildlife, sold (for any purpose), in a fresh-food market? | Yes |
| Is the record in English? | Yes |
| Is the record primary research? | Yes |

Level 2 Screening on full record

| Question | Inclusion value |
| --- | --- |
| Is the full text available? | Yes |
| Is the record in English? | Yes |
| Does the record include information about live terrestrial wildlife, sold (for any purpose), in a fresh-food market? | Yes |
| Is the record primary research? | Yes |

Level 3 Data charting:

ID

BACKGROUND

Publication Year

Study years (if range format: 2019-2020. If multiple periods, list each year: 2018, 2020)

Study country

Study region (OR towns if region NOT listed)

What are the drivers for this study? Choice = Understanding wildlife trade, understanding disease, other.

If 'other' drivers or the drivers are not clear, give details/paste in relevant text rom record.

Is this a multi-study record (e.g. a thesis with two relevant chapters)?

If this is a multi-study record, identify your line responses (e.g. Line 1 = Chapter 3, Line 2 = chapter 4).

MARKETS

What type of market is it?

If 'other' market type, describe this.

Is there information about the market physical structure?

If yes, give information about market structure.

Is there information about market frequency?

If there is information about market frequency, give details.

List the types of people associated with the market (e.g. vendors, collectors, hunters).

Is there information about activities associated with the wildlife? e.g. slaughter of wildlife in the market, caging animals for transport, preparation

If there is information about activities at the market, give details (e.g. type of person involved and the activity).

Are hygiene practices mentioned?

If hygiene practices are mentioned, give details.

Are temporal changes in the sale of wildlife mentioned (e.g. frequency of market, diversity or number of species, changes in reasons for sale)?

If temporal changes are mentioned, give details.

ANIMALS

Does the record list species of wildlife that were sold LIVE?

If live species were mentioned, list their names (common and Latin).

Select classes of WILDLIFE sold at the market (LIVE or DEAD).

List all orders in each class of WILDLIFE sold at the market (see attached PDF).

Is there information about the source of the LIVE wildlife (e.g. local, imported)?

If there is information about the source, give details.

Does the record indicate the volume of LIVE wildlife sold at the market (e.g. the researchers saw one live pangolin; or, 5.2% of wildlife were sold live)

If the record indicates the volume of LIVE wildlife, give details.

Does the record indicate the TOTAL volume of wildlife (dead or alive) sold at the market? (Not just the live wildlife)

If the record indicates the volume of TOTAL wildlife, give details.

Does the record indicate the purpose of the wildlife sale?

If the record indicates the purpose of the wildlife sale, give details.

Does the record indicate the condition of the LIVE wildlife (e.g. stress, disease, health, injury)?

If the record indicates the condition of the LIVE wildlife, give details.

Does the record give information about the housing of the live wildlife?

If the record gives information about housing, give details.

Does the record mention other non-wildlife species at the market (e.g. vermin, domestic species)?

If the record mentions other species, list them.

Does the record mention contact between LIVE wildlife and other LIVE species at the market (direct between live animals, or indirect, e.g. post-slaughter)?

If the record mentions contact between live wildlife and other live animals, give details.

RISKS

Does the record identify diseases associated with the market (in the results)?

If diseases were identified, give details (disease and location and method of diagnosis/identification).

Does the record identify microbes (pathogen or not) associated with the market (in the results)?

If microbes were identified, give details (microbe and location and method of identification).

If microbes were identified, give details of measures of frequency.

Were any other risks associated with human and animals health identified in the record?

If other risk were identified, give details.
