## Supplementary Material 2 for "Live wildlife trade in markets – a scoping review to inform risk assessment of emerging infectious diseases"

| ID | Title | Authors | Publication | PubYea | Study_Years1 | Study_Years2 | Country_1 | Country_Study | Global_region | Global_region2 | Global_region3 | Region | Driver_study | Driver_other |
|---|---|---|---|---|---|---|---|---|---|---|---|---|---|---|
| Akani2015 | Are mammal communities occurring at a regional scale reliably represented in "hub" bushmeat markets? A case study with Bayelsa State (Niger Delta, Nigeria). | Akani, G. C., Amadi, N., Eniang, E. A., Luiselli, L., & Petrozzi, F. | Folia Zoologica | 2015 | 2013-2014 | 2013, 2014 | Nigeria | Nigeria | Africa | Africa | Africa | Bayelsa State, Niger Delta | wildlife trade | NA |
| Cronin2015 | Long-Term Urban Market Dynamics Reveal Increased Bushmeat Carcass Volume despite Economic Growth and Proactive Environmental Legislation on Bioko Island, Equatorial Guinea | Cronin, D.T., Woloszynek, S., Morra, W.A., Honarvar, S., Linder, J.M., Gonder, M.K., O'Connor M.P., Hearn G.W. | Plos One | 2015 | 1997-2010 | 1997, 1998, 1999, 2000, 2001, 2002, 2003, 2004, 2005, 2006, 2007, 2008, 2009, 2010 | Equatorial Guinea | Equatorial Guinea | Africa | Africa | Africa | Bioko Island | wildlife trade | NA |
| DCruze2020 | Snake oil and pangolin scales: insights into wild animal use at "Marché des Fétiches" traditional medicine market, Togo | D'Cruze, N., Assou, D., Coulthard, E., Norrey, J., Megson, D., MacDonald, D.W., Harrington, L.A., Ronfot, D., Segniagbeto, G.H., Auliya, M. | Nature Conservation | 2020 | 2018, 2019 | 2018, 2019 | Togo | Togo | Africa | Africa | Africa | Lome | wildlife trade | NA |
| Djagoun2018 | Trade in primate species for medicinal purposes: implications for conservation. | Sogbohossou, E. A., Kassa, B., Akpona, H. A., Amahowe, I. O., Djagoun, J., & Sinsin, B. | TRAFFIC Bulletin | 2018 | 2012 | 2012 | Benin | Benin | Africa | Africa | Africa | Southern Benin | wildlife trade | NA |

| ID | MarketType | Market_cleaned | MarketType_other | Structur | Structure_detail | Frequenc | Freq_detail | People | Activi | Activity_detail |
|---|---|---|---|---|---|---|---|---|---|---|
| Akani2015 | Bushmeat | Bushmeat | NA | Yes | It is situated between five forested reserves of the Niger Delta. The market is located on the shore of the River Nun. Thus, hunters easily transport their bounties by rivercraft. Swali market is also accessible by good roads, recently constructed by the Bayelsa State government. | No | NA | Hunters, bushmeat dealers | No | NA |
| Cronin2015 | Bushmeat | Bushmeat | NA | No | NA | Yes | The market operated year-round, Monday-Saturday, with occasional Sunday activity until approximately May 2010, after which the market sporadically closed on Tuesdays for | Intermediaries (bringing animals to market), vendors | No | NA |
| DCruze2020 | Fetish | Fetish | NA | Yes | Stalls | No | NA | Vendors | No | NA |
| Djagoun2018 | Traditional medicine market | Traditional medicine | NA | Yes | Market in city of Porto-Novo is close to wetlands (Ouémé valley, Hlan river) part of Ramsar site 1,017, and close to Nigeria. Markets in the city of Cotonou [illegible] close to wetlands (Ramsar site 1,018) and close to Togo. The market in the city of Cotonou is the biggest market in Benin. Some markets were "outdoor markets" | Yes | Some markets were occasional markets (markets set up for special events) | animal-based medicine traders, traditional healers, customers | No | NA |

| ID | Illegal_activit | Illegal_detail | Hygien | Hygiene_detail | TemporalChang | TemporalChange_detail |
|---|---|---|---|---|---|---|
| Akani2015 | Yes | Species that officially are protected by Nigerian federal laws were regularly traded. "Local taboos were also unlikely to affect the trade at Swali market, because this marketplace collated animals from a suite of different forest sites where hunting was performed by different ethnic groups." | No | NA | Yes | The collected data suggest a heavier trade during the wet months... the numbers of traded carcasses were higher during the wet season at a marginally significant level (χ2 = 30.9, df = 20, P = 0.0501). In addition, these inter-seasonal differences were highly significant in terms of traded biomass (higher by wet season; χ2 = 390.2, df = 20, P < 0.0001) |
| Cronin2015 | Yes | Trade in species from legally protected areas | Yes | From 2010, closed occasionally Tuesday for cleaning | Yes | Market growth was consistent over time, as the mean number of carcasses per market day increased progressively over the three periods: 29.96 (SD 4.94), 50.46 (SD 16.65), and 91.86 (SD 12.11) carcasses/day, respectively. |
| DCruze2020 | Yes | Over 20% of species should have been protected from trade by CITES. Local laws are difficult to assess. | No | NA | Yes | Fewer pangolins than 5 years ago |
| Djagoun2018 | Yes | Several species were illegal to be traded inBenin. Also, several must have been illegally reported. 'Corrupt security agents and porous borders in Benin are suspected to be the main drivers of likely illegal trade in contravention of CITES.' | No | NA | No | NA |

| ID | ListLive_Y | ListLive_Cleaned | ListLive_Cleaned_Genus | Classes_LiveDead | Orders_cleaned_LiveDead |
|---|---|---|---|---|---|
| Akani2015 | Yes | Mona monkey (Cercopithecus mona), Sclaters guenon (Cercopithecus sclateri), Greater spot nosed monkey (Cercopithecus nictitans) | Pangolin (Phataginus) | Mammalia, Reptilia | Primates, Cetartiodactyla, Rodentia, Pholidota, Carnivora, Squamata |
| Cronin2015 | No | NA | | Aves, Mammalia, Reptilia | Crocodilia, Squamata, Testudines, Rodentia, Primates, Pholidota, Carnivora, Artiodactyla, Bucerotiformes, Musophagiformes, Accipitriformes |
| DCruze2020 | Yes | Ball python (Python regius), Chameleon (Chamaeleo chamaeleon), Vulture (Gypohierax angolensis) | Crocodile (Crocodylus), Cane rat (Thryonomys), Monitor lizard (Varanus) | Mammalia, Aves, Reptilia, Amphibia | Artiodactylae, Primates, Chiroptera, Carnivora, Squamata, Crocodilia, Accipitriformes, Proboscidea, Amphibia, Rodentia, Strigiformes, Pholidota, Psittaciformes, Galliformes, Passeriformes, Testudines |
| Djagoun2018 | Yes | Senegal bushbaby (Galago senegalensis) | | Mammalia | Primates |

| ID | Sourc | Source_detail | Farm | Local_TransB | VolumeLiv | VolumeLive_detail | VolumeTota | VolumeTotal_detail |
|---|---|---|---|---|---|---|---|---|
| Akani2015 | Yes | Most of the hunters catch the animals from the thick seasonally inundated swamp forests that are situated within 15 km radius from Swali market, i.e. Emeyal, Otuasega, Elebeli, Famgbe, Bebelebiri, Kolo Creek and Ogbia areas. It is also used sometimes for sale of remotely captured animals. For instance, we recorded a few bushmeat items belonging to species that do not occur in the Niger Delta, such as the puff adder, Bitis arietans. [Note - source of bushmeat not necessarily and specifically LIVE animals] | NA | Local | Yes | 12 Cercopithecus mona, 2 Cercopithecus sclateri, 8 Cercopithecus nictitans, and 5 pangolins were alive at the examination time | Yes | 9365.73 kg |
| Cronin2015 | Yes | Most carcasses are from the island, but imported carcasses increasing. | NA | Local, Exotic | Yes | Most carcasses (81.9%) were fresh (recently killed) when they arrived in the market and only 12.6% were smoked. Few animals (5.1%) reached the market alive. | Yes | 196,882 carcasses over 3,758 market days in the Malabo market between October 1997 and September 2010. |
| DCruze2020 | Yes | Local and neighbouring countries, e.g. Benin and Nigeria | NA | Local, Exotic | Yes | The most frequently cited type of product traded was a live animal (n = 43, 33%). | No | NA |
| Djagoun2018 | Yes | The principal origin of the primate products displayed in the markets was Nigeria, but several countries of West and Central Africa also feed the trade in primate products on sale in Benin According to information received from traders, most of the primates and other animals sold in Benin markets come from the country's national parks or gazetted forests. However, although most of the primates on sale occur in Benin, the authors discovered that traders also imported them from other African countries. Primates in the traditional medicine markets in Benin primarily come from Nigeria (34%), followed by Burkina Faso (16%), Ghana (16%), Niger (11%), Togo (8%), Gabon (6.6%), Côte d'Ivoire (4.8%) and Mali (3%). Central African countries as a source of supply, including Congo and Cameroon, were cited only by 2.5% and 2% of traders respectively. [Note this does not specifically indicate that these were sources of LIVE animals] | NA | Local, Exotic | No | NA | No | NA |

| ID | Purpos | Purpose_detail | Purpose1 | Conditio | Condition_detail | Housin | Housing_detail | SpeciesOthe | SpeciesOther_list |
|---|---|---|---|---|---|---|---|---|---|
| Akani2015 | Yes | Human consumption | Food | No | NA | No | NA | No | NA |
| Cronin2015 | Yes | Bushmeat | Food | No | NA | No | NA | No | NA |
| DCruze2020 | Yes | Medicine, spiritual use | Medicine, Spiritual | Yes | The housing listed 'clearly compromised their health and wellbeing'. | Yes | small barren dirty cages, cloth bags and plastic buckets' | No | NA |
| Djagoun2018 | Yes | All species are believed to have both medicinal and magical values, however their use for magical purposes predominates. | Medicine, Spiritual | No | NA | No | NA | No | NA |

| ID | ContactLiv | ContactLive_detail | DiseaseId | DiseaseId_detail | Microbeid | MicrobeId_detail | DetectionMethods | OtherRisk | OtherRisks_detail |
|---|---|---|---|---|---|---|---|---|---|
| Akani2015 | No | NA | No | NA | No | NA | NA | No | NA |
| Cronin2015 | No | NA | No | NA | No | NA | NA | No | NA |
| DCruze2020 | No | NA | No | NA | No | NA | NA | No | NA |
| Djagoun2018 | No | NA | No | NA | No | NA | NA | Yes | Though no direct diseases or pathogens were specifically identified in the markets from this study, the article does include the following passage raising the possibility of disease risk. "Several species of monkeys have been identified as harboring infectious diseases transmissible to humans, with potentially grave consequences (Still, 2003). Organs and various tissues, including bones and bile, can be a source of Salmonella infection, causing chronic diarrhoea and endotoxic shock, but also the recently emerging disease Ebola that can inter alia be caused by the consumption of infected primate parts. The possibility of transmission of other serious and widespread zoonoses such as tuberculosis or rabies should be considered whenever animal tissues from unknown sources are handled and used as remedies (Alves and Rosa, 2007)." |
