## Supplementary Material 3 for "Live wildlife trade in markets – a scoping review to inform risk assessment of emerging infectious diseases"

V.J. Brookes^1, 2^, O. Wismandanu^3^, E. Sudarnika^4^, J.A. Roby^2, 5^, L. Hayes^1,2^, M.P. Ward^6^, C. Basri^4^, H. Wibawa^7^, J. Davis^8^, D. Indrawan^9^, J. Manyweathers^1, 2,^ W.S. Nugroho^10^, S. Windria^11^, M. Hernandez-Jover^1, 2^

^1^ School of Animal and Veterinary Sciences, Charles Sturt University, Australia

^2^ Graham Centre for Agricultural Innovation, (NSW Dept. of Primary Industries and Charles Sturt University), Australia

^3^ Veterinary Medicine Study Program, Faculty of Medicine, Padjadjaran University, Indonesia

^4^ Faculty of Veterinary Medicine, IPB (Institut Pertanian Bogor) University, Indonesia

^5^ School of Biomedical Sciences, Faculty of Science, Charles Sturt University, Australia

^6^ Sydney School of Veterinary Science, The University of Sydney, Australia

^7^ Disease Investigation Centre Wates, Directorate General of Livestock and Animal Health Services, Ministry of Agriculture of Indonesia

^8^ Australian Department of Agriculture, Water and the Environment, Canberra, Australia

^9^ School of Business, IPB University, Indonesia

^10^ Faculty of Veterinary Medicine, Universitas Gadjah Mada, Indonesia

^11^ Department of Biomedical Sciences, Division of Microbiology, Veterinary Medicine Study Program, Faculty of Medicine, Padjadjaran University, Indonesia

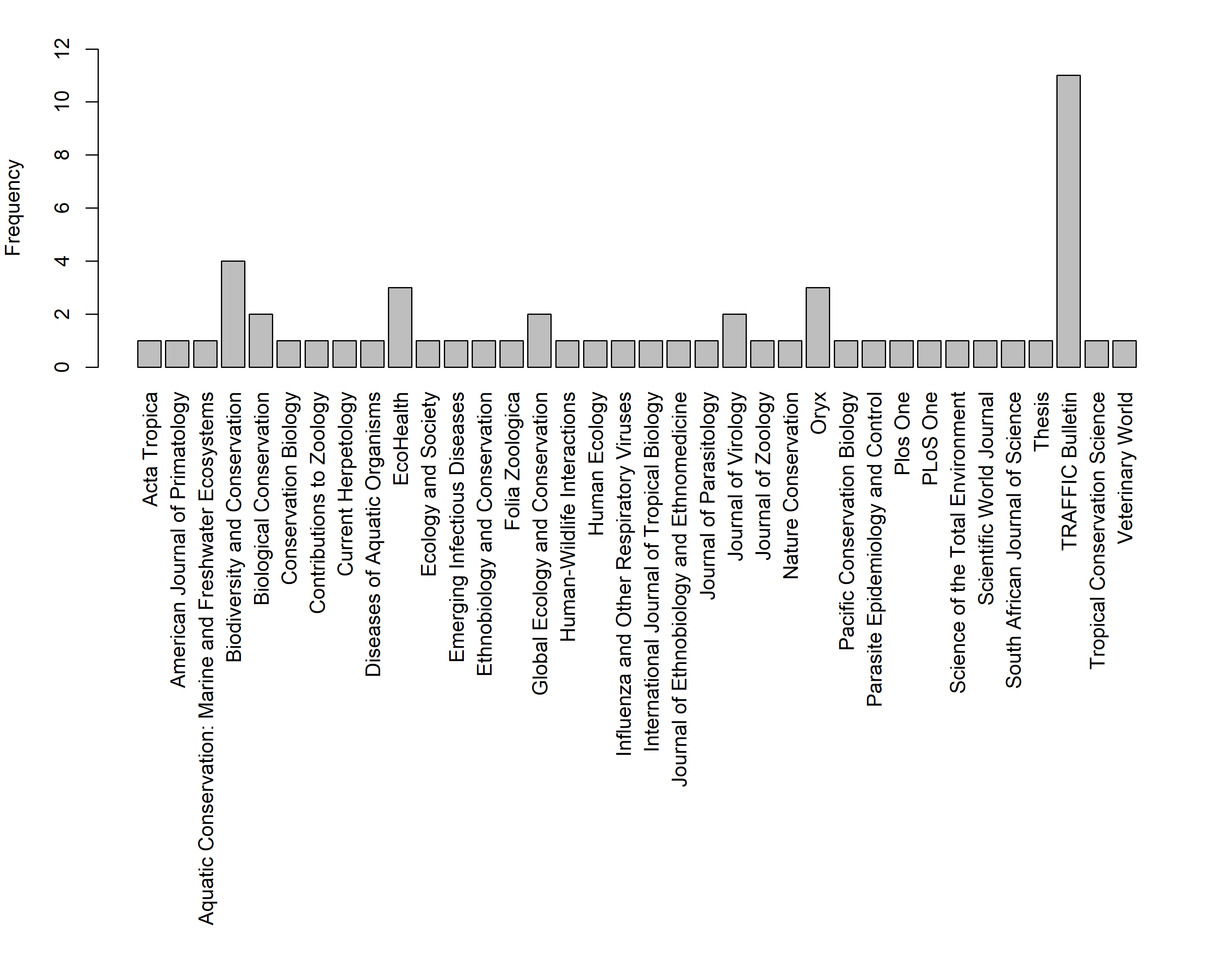

Figure S1 Frequency of publications in peer-reviewed journals in a scoping review of research associated with live, terrestrial, vertebrate wildlife sold for any purpose at markets likely to sell food.

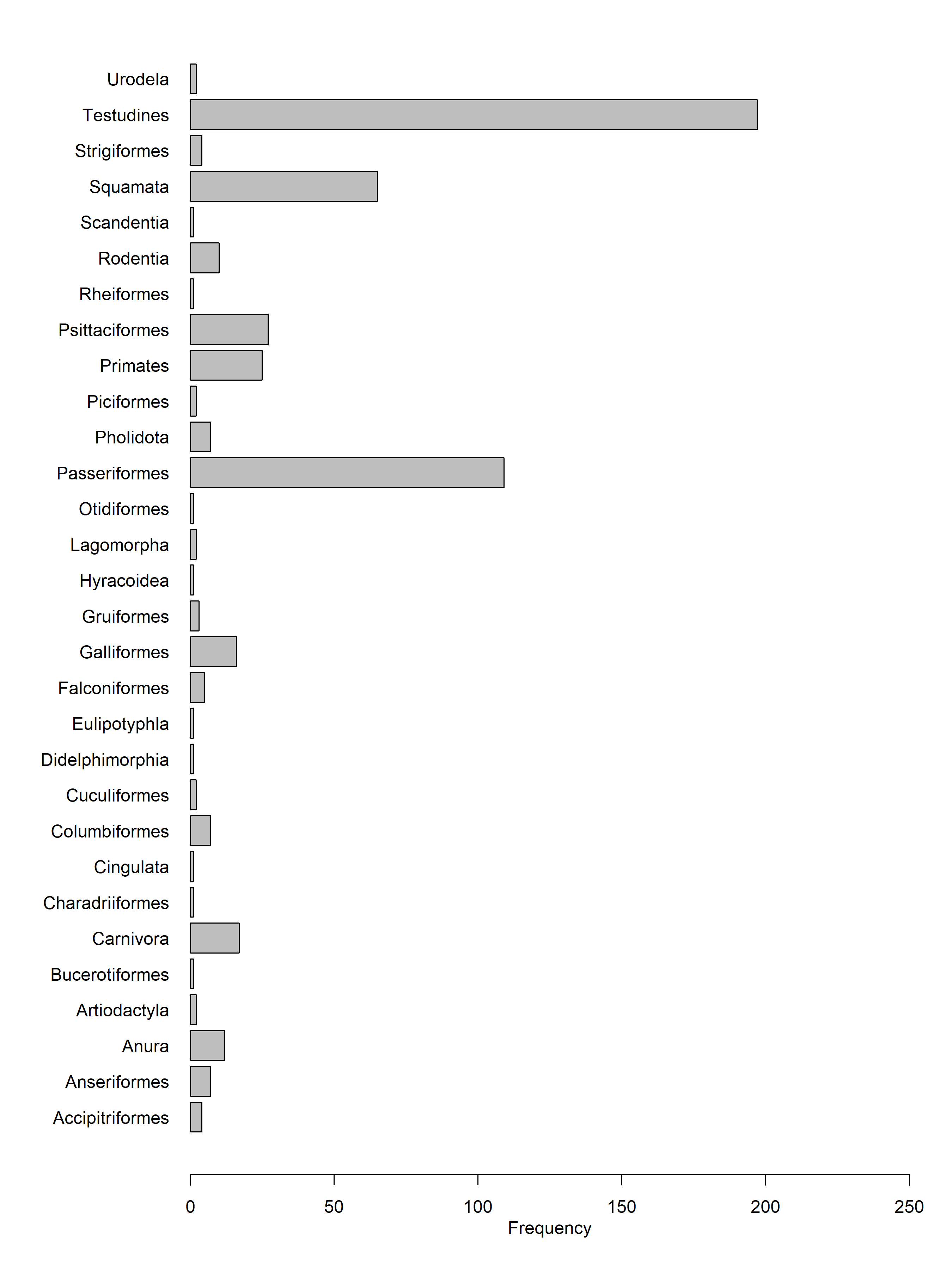

Figure S2 Frequency of orders of live wildlife reported in markets in a scoping review of research associated with live, terrestrial, vertebrate wildlife sold for any purpose at markets likely to sell food.

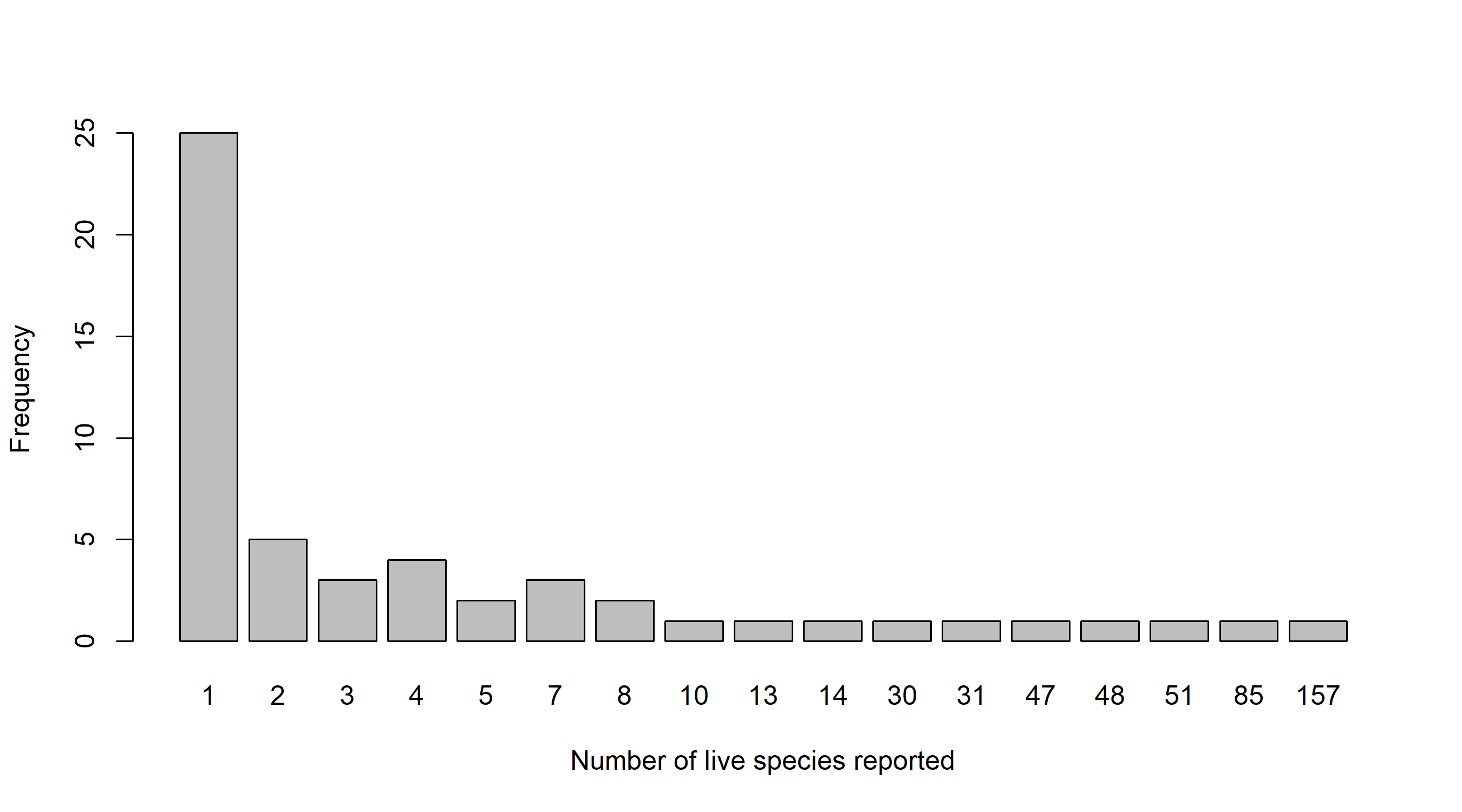

Figure S3 Frequency of records reporting numbers of live species of wildlife reported in markets in a scoping review of research associated with live, terrestrial, vertebrate wildlife sold for any purpose at markets likely to sell food.

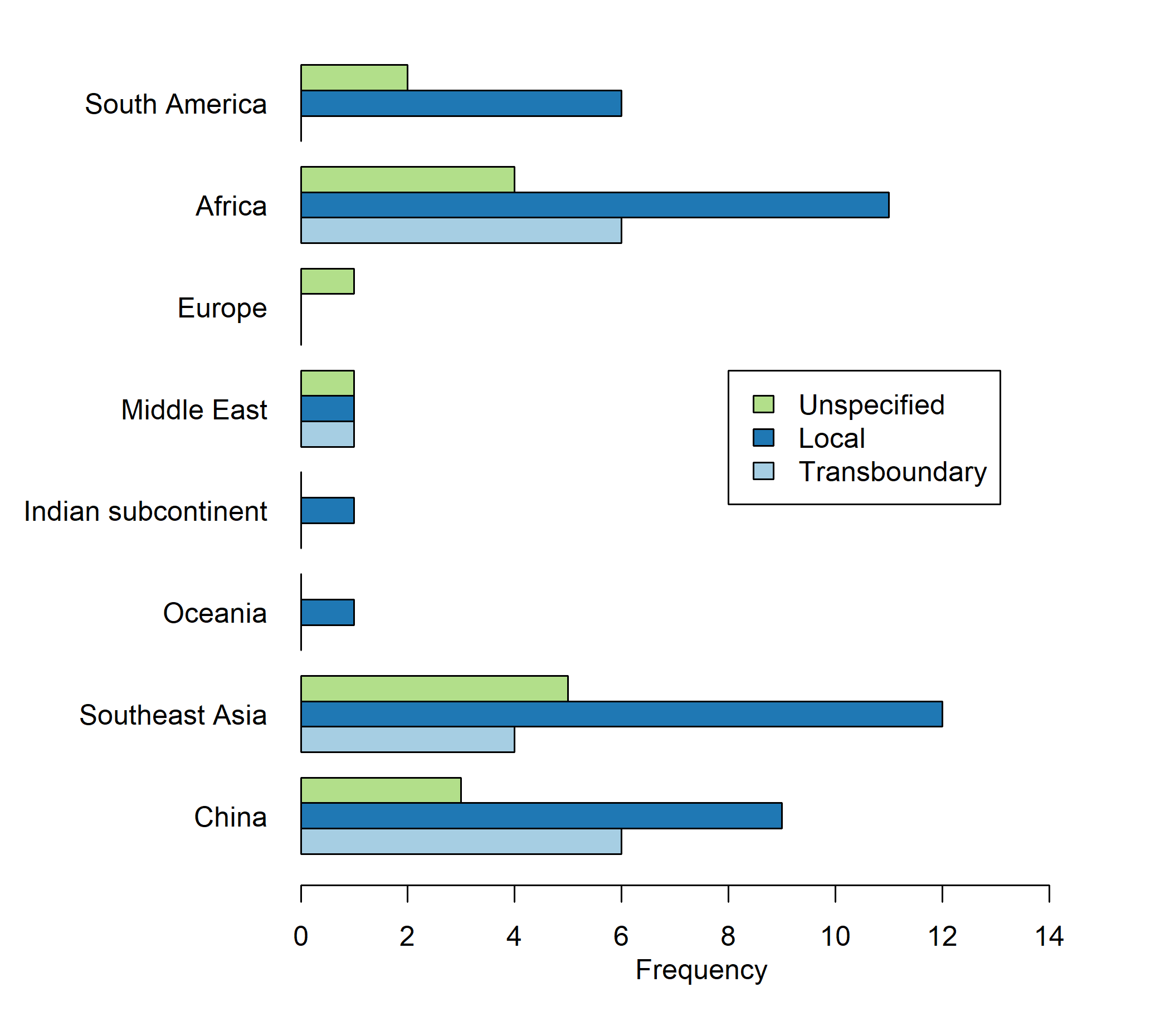

Figure S4 Frequency of records reporting origin of live species of wildlife reported in markets in a scoping review of research associated with live, terrestrial, vertebrate wildlife sold for any purpose at markets likely to sell food.

Table S1 List of live, vertebrate, terrestrial wildlife species, reported in markets in a scoping review of research associated with live, terrestrial, vertebrate wildlife sold for any purpose at markets likely to sell food.

| Region | Class | Order | Common name | Latin | Frequency |
| --- | --- | --- | --- | --- | --- |
| Africa | Aves | Accipitriformes | Palm-nut vulture | *Gypohierax angolensis* | 1 |
|  |  | Falconiformes | Common kestrel | *Falco tinnunculus* | 1 |
|  | Mammalia | Pholidota | Giant pangolin | *Smutsia gigantea* | 1 |
|  |  |  | Tree pangolin | *Manis tricuspis* | 2 |
|  |  |  | Tree pangolin | *Phataginus tricuspis* | 1 |
|  |  | Primates | Chimpanzee | *Pan troglodytes* | 1 |
|  |  |  | Greater spot nosed monkey | *Cercopithecus nictitans* | 1 |
|  |  |  | Mona monkey | *Cercopithecus mona* | 1 |
|  |  |  | Patas monkey | *Erythrocebus patas* | 1 |
|  |  |  | Sclaters guenon | *Cercopithecus sclateri* | 1 |
|  |  |  | Senegal bushbaby | *Galago senegalensis* | 1 |
|  | Reptilia | Squamata | Ball python | *Python regius* | 2 |
|  |  |  | Bell’s dabb lizard | *Uromastryx acanthinura* | 1 |
|  |  |  | Common chameleon | *Chamaeleo chamaeleon* | 3 |
|  |  |  | Flap-necked chameleon | *Chamaeleo dilepis* | 1 |
|  |  |  | Desert monitor lizard | *Varanus griseus* | 1 |
|  |  |  | Egyptian cobra | *Naja haje* | 1 |
|  |  |  | Nile monitor | *Varanus niloticus* | 1 |
|  |  |  | Savannah monitor | *Varanus exanthematicus* | 1 |
|  |  |  | Veiled chameleon | *Chamaeleo calyptratus* | 1 |
|  |  | Testudines | Spur thighed tortoise | *Testudo graeca* | 1 |
| China | Amphibia | Anura | American bullfrog | *Rana catesbeiana* | 2 |
|  |  |  | Boulenger’s toad | *Rana boulengeri* | 1 |
|  |  |  | Dark spotted frog | *Rana nigromaculata* | 1 |
|  |  |  | Tiger frog | *Rana tigerina* | 1 |
|  |  | Urodela | Black knobby newt | *Echinotriton asperrimus* | 1 |
|  |  |  | Chinese giant salamander | *Andrias davidianus* | 1 |
|  | Aves | Accipitriformes | Black kite | *Milvus migrans* | 1 |
|  |  | Anseriformes | Mallard | *Anas platyrhynchos* | 1 |
|  |  |  | Mandarin duck | *Aix galericulata* | 1 |
|  |  | Columbiformes | Oriental turtle dove | *Streptopelia orientalis* | 1 |
|  |  |  | Spotted dove | *Streptopelia chinensis* | 1 |
|  |  | Cuculiformes | Greater coucal | *Centropus sinensis* | 1 |
|  |  | Falconiformes | Eurasian sparrowhawk | *Accipiter nisus* | 1 |
|  |  | Galliformes | Chinese francolin | *Francolinus pintadeanus* | 1 |
|  |  |  | Common pheasant | *Phasianus colchicus* | 2 |
|  |  |  | Elliot’s pheasant | *Syrmaticus ellioti* | 1 |
|  |  |  | Golden pheasant | *Chrysolophus pictus* | 1 |
|  |  |  | Grey peacock pheasant | *Polyplectron bicalcaratum* | 1 |
|  |  |  | Hume’s pheasant | *Syrmaticus humiae* | 1 |
|  |  |  | Lady amherst’s pheasant | *Chrysolophus amherstiae* | 1 |
|  |  | Galliformes | Silver pheasant | *Lophura nycthemera* | 1 |
|  |  |  | Temminck’s tragopan | *Tragopan temminckii* | 1 |
|  |  |  | White breasted waterhen | *Amaurornis phoenicurus* | 1 |
|  |  | Passeriformes | Eurasian lark | *Alauda arvensis* | 1 |
|  |  |  | Black billed magpie | *Pica pica* | 1 |
|  |  |  | Black throated laughing thrush | *Garrulax chinensis* | 1 |
|  |  |  | Chestnut-flanked white-eye | *Zosterops erythropleurus* | 1 |
|  |  |  | Common hill mynah | *Gracula religiosa* | 2 |
|  |  |  | Crimson sunbird | *Aethopyga siparaja* | 1 |
|  |  |  | Horned lark | *Eremophila alpestris* | 1 |
|  |  |  | Hwamei | *Garrulax canorus* | 1 |
|  |  |  | Japanese white-eye | *Zosterops japonicus* | 1 |
|  |  |  | Marsh tit | *Poecile palustris* | 1 |
|  |  |  | Masked laughing thrush | *Garrulax perspicillatus* | 1 |
|  |  |  | Mongolian lark | *Melanocorypha mongolica* | 1 |
|  |  |  | Red billed leiothrix | *Leiothrix lutea* | 1 |
|  |  |  | Silver eared mesia | *Leiothrix argentauris* | 1 |
|  |  |  | White browed laughing thrush | *Garrulax sannio* | 1 |
|  |  |  | White throated laughing thrush | *Garrulax albogularis* | 1 |
|  |  |  | Willow tit | *Poecile montanus* | 1 |
|  |  | Psittaciformes | Derbyan parakeet | *Psittacula derbiana* | 1 |
|  |  |  | Plum headed parakeet | *Psittacula cyanocephala* | 1 |
|  |  |  | Red breasted parakeet | *Psittacula alexamdri* | 1 |
|  |  |  | Rose ringed parakeet | *Psittacula krameri* | 1 |
|  |  | Strigiformes | African grass owl | *Tyto capensis* | 1 |
|  |  |  | Eurasian eagle owl | *Bubo bubo* | 2 |
|  |  |  | Long eared owl | *Asio otus* | 1 |
|  | Mammalia | Carnivora | Asian leopard cat | *Prionailurus bengalensis* | 1 |
|  |  |  | Chinese ferret badger | *Melogale moschata* | 1 |
|  |  |  | Crab eating mongoose | *Herpestes urva* | 1 |
|  |  |  | Large indian civet | *Viverra zibetha* | 1 |
|  |  |  | Leopard cat | *Felis bengalensis* | 1 |
|  |  |  | Lesser indian civet | *Viverricula indica* | 2 |
|  |  |  | Masked palm civet | *Paguma larvata* | 3 |
|  |  |  | Siberian weasel | *Mustela sibirica* | 1 |
|  |  |  | Yellow bellied weasel | *Mustela kathiah* | 1 |
|  |  | Eulipotyphla | European hedgehog | *Erinaceus europaeus* | 1 |
|  |  | Lagomorpha | Chinese hare | *Lepus sinensis* | 1 |
|  |  | Pholidota | Chinese pangolin | *Manis pentadactyla* | 1 |
|  |  | Primates | Assam macaque | *Macaca assamensis* | 2 |
|  |  |  | Black gibbon | *Hylobates concolor* | 1 |
|  |  |  | Francois' leaf monkey | *Trachypithecus francoisi* | 1 |
|  |  |  | Hoolock gibbon | *Hylobates hoolock* | 1 |
|  |  |  | Lesser slow loris | *Nycticebus pygmaeus* | 1 |
|  |  |  | Long tailed macaque | *Macaca fascicularis* | 1 |
|  |  |  | Pig tailed macaque | *Macaca nemestrina* | 2 |
|  |  | Primates | Rhesus macaque | *Macaca mulatta* | 2 |
|  |  |  | Sunda slow loris | *Nycticebus coucang* | 2 |
|  |  |  | Stump tailed macaque | *Macaca arctoides* | 2 |
|  |  |  | Tonkin snub nosed monkey | *Pygathrix avunculus* | 1 |
|  |  | Rodentia | Chinese bamboo rat | *Rhizomys sinensis* | 2 |
|  |  |  | Chinese porcupine | *Hystrix hodgsoni* | 1 |
|  |  |  | Hoary bamboo rat | *Rhizomys pruinosus* | 1 |
|  |  |  | Pallas's squirrel | *Callosciurus erythraeus* | 1 |
|  |  | Scandentia | Malaysian treeshrew | *Tupaia glis* | 1 |
|  | Reptilia | Squamata | Asian cobra | *Naja naja* | 3 |
|  |  |  | Asian rock python | *Python molurus* | 2 |
|  |  |  | Banded krait | *Bungarus fasciatus* | 3 |
|  |  |  | Beauty rat snake | *Elaphe taeniura* | 1 |
|  |  |  | Brown spotted pit viper | *Trimeresurus mucrosquamatus* | 1 |
|  |  |  | Buff striped keelback | *Amphiesma stolata* | 1 |
|  |  |  | Cantor’s rat snake | *Ptyas dhumnades* | 1 |
|  |  |  | Cantor’s rat snake | *Pytas dhumnades* | 1 |
|  |  |  | Chinese crocodile lizard | *Shinisaurus crocodilurus* | 1 |
|  |  |  | Chinese rat snake | *Pytas korros* | 3 |
|  |  |  | Chinese water dragon | *Physignathus cocincinus* | 1 |
|  |  |  | Green snake | *Opheodrys major* | 1 |
|  |  |  | Hart's glass lizard | *Ophisaurus harti* | 1 |
|  |  |  | King cobra | *Ophiophagus hannah* | 2 |
|  |  |  | King rat snake | *Elaphe carinata* | 1 |
|  |  |  | Many banded krait | *Bungarus multicinctus* | 2 |
|  |  |  | Moellendorf's rat snake | *Elaphe moellendorffi* | 1 |
|  |  |  | Oriental garden lizard | *Calotes versicolor* | 1 |
|  |  |  | Oriental long tailed lizard | *Takydromus sexlineatus* | 1 |
|  |  |  | Oriental rat snake | *Ptyas mucosus* | 3 |
|  |  |  | Pit viper | *Deinagkistrodon acutus* | 2 |
|  |  |  | Rat snake | *Elaphe radiata* | 3 |
|  |  |  | Red keelback | *Macropisthodon rudis* | 1 |
|  |  |  | Rice paddy snake | *Hypsiscopus plumbea* | 1 |
|  |  |  | Russell’s viper | *Vipera russelli* | 1 |
|  |  |  | Slender sea snake | *Microcephalophis gracilis* | 1 |
|  |  |  | Stejneger's pit viper | *Trimeresurus stejnegeri* | 1 |
|  |  |  | Thin ground snake | *Elaphe taeniurus* | 1 |
|  |  |  | Tokay gecko | *Gecko gecko* | 2 |
|  |  |  | Water monitor | *Varanus salvator* | 2 |
|  |  | Testudines |  | *Aspideretes hurum* | 1 |
|  |  |  |  | *Batagur baska* | 1 |
|  |  |  |  | *Callagur borneoensis* | 1 |
|  |  |  |  | *Chelodina novaeguinae* | 1 |
|  |  |  |  | *Chelodina parkeri* | 1 |
|  |  |  |  | *Chelodina siebenrocki* | 1 |
|  |  |  |  | *Chelonia mydas* | 1 |
|  |  |  |  | *Chelydra rossignonii* | 1 |
|  |  |  |  | *Chelydra serpentina* | 1 |
|  |  |  |  | *Chersina angulata* | 1 |
|  |  |  |  | *Chinemys nigricans* | 1 |
|  |  |  |  | *Chitra chitra* | 1 |
|  |  |  |  | *Chitra indica* | 1 |
|  |  |  |  | *Claudius angustatus* | 1 |
|  |  |  |  | *Clemmys guttata* | 1 |
|  |  |  |  | *Clemmys insculpta* | 1 |
|  |  |  |  | *Clemmys marmorata* | 1 |
|  |  |  |  | *Clemmys muhlenbergii* | 1 |
|  |  |  |  | *Cuora aurocapitata* | 1 |
|  |  |  |  | *Cuora mccordi* | 1 |
|  |  |  |  | *Cuora pani* | 1 |
|  |  |  |  | *Cyclanorbis elegans* | 1 |
|  |  |  |  | *Cyclemys tcheponensis* | 1 |
|  |  |  |  | *Cycloderma frenatum* | 1 |
|  |  |  |  | *Deirochelys reticularia* | 1 |
|  |  |  |  | *Dogania subplana* | 1 |
|  |  |  |  | *Emydoidea blandingii* | 1 |
|  |  |  |  | *Emydura subglobosa* | 1 |
|  |  |  |  | *Emys orbicularis* | 1 |
|  |  |  |  | *Geochelone carbonaria* | 1 |
|  |  |  |  | *Geochelone chilensis* | 1 |
|  |  |  |  | *Geochelone denticulata* | 1 |
|  |  |  |  | *Geochelone elegans* | 1 |
|  |  |  |  | *Geochelone gigantea* | 1 |
|  |  |  |  | *Geochelone pardalis* | 1 |
|  |  |  |  | *Geochelone platynota* | 1 |
|  |  |  |  | *Geochelone radiata* | 1 |
|  |  |  |  | *Geochelone sulcata* | 1 |
|  |  |  |  | *Geoemyda japonica* | 1 |
|  |  |  |  | *Gopherus flavomarginatus* | 1 |
|  |  |  |  | *Graptemys barbouri* | 1 |
|  |  |  |  | *Graptemys caglei* | 1 |
|  |  |  |  | *Graptemys ernsti* | 1 |
|  |  |  |  | *Graptemys geographica* | 1 |
|  |  |  |  | *Graptemys gibbonsi* | 1 |
|  |  |  |  | *Graptemys nigrinoda* | 1 |
|  |  |  |  | *Graptemys pseudogeographica* | 1 |
|  |  |  |  | *Graptemys pulchra* | 1 |
|  |  |  |  | *Graptemys versa* | 1 |
|  |  |  |  | *Heosemys grandis* | 1 |
|  |  |  |  | *Heosemys spinosa* | 1 |
|  |  |  |  | *Hieremys annandalii* | 1 |
|  |  |  |  | *Homopus areolatus* | 1 |
|  |  |  |  | *Homopus signatus* | 1 |
|  |  |  |  | *Hydromedusa maximiliani* | 1 |
|  |  |  |  | *Hydromedusa tectifera* | 1 |
|  |  |  |  | *Indotestudo forstensii* | 1 |
|  |  |  |  | *Kachuga dhongoka* | 1 |
|  |  |  |  | *Kachuga kachuga* | 1 |
|  |  |  |  | *Kachuga smithii* | 1 |
|  |  |  |  | *Kachuga sylhetensis* | 1 |
|  |  |  |  | *Kachuga tecta* | 1 |
|  |  |  |  | *Kachuga tentoria* | 1 |
|  |  |  |  | *Kachuga trivittata* | 1 |
|  |  |  |  | *Kinixys belliana* | 1 |
|  |  |  |  | *Kinixys erosa* | 1 |
|  |  |  |  | *Kinixys homeana* | 1 |
|  |  |  |  | *Kinixys spekii* | 1 |
|  |  |  |  | *Kinosternon flavescens* | 1 |
|  |  |  |  | *Kinosternon leucostomum* | 1 |
|  |  |  |  | *Kinosternon scorpioides* | 1 |
|  |  |  |  | *Kinosternon subrubrum* | 1 |
|  |  |  |  | *Kinsternon sonoriense* | 1 |
|  |  |  |  | *Leucocephalon yuwonoi* | 1 |
|  |  |  |  | *Malaclemys terrapin* | 1 |
|  |  |  |  | *Malacochersus tornieri* | 1 |
|  |  |  |  | *Manouria impressa* | 2 |
|  |  |  |  | *Mauremys caspica* | 1 |
|  |  |  |  | *Mauremys japonica* | 1 |
|  |  |  |  | *Melanochelys tricarinata* | 1 |
|  |  |  |  | *Morenia ocellata* | 1 |
|  |  |  |  | *Notochelys platynota* | 1 |
|  |  |  |  | *Ocadia philippeni* | 1 |
|  |  |  |  | *Orlitia borneensis* | 1 |
|  |  |  |  | *Pelochelys cantorii* | 1 |
|  |  |  |  | *Pelusios subniger* | 1 |
|  |  |  |  | *Phrynops geoffroanus* | 1 |
|  |  |  |  | *Phrynops hilairii* | 1 |
|  |  |  |  | *Phrynops williamsi* | 1 |
|  |  |  |  | *Platemys platycephala* | 1 |
|  |  |  |  | *Podocnemis expansa* | 1 |
|  |  |  |  | *Podocnemis sextuberculata* | 1 |
|  |  |  |  | *Psammobates oculiferus* | 1 |
|  |  |  |  | *Psammobates tentorius* | 1 |
|  |  |  |  | *Pseudemys nelsoni* | 1 |
|  |  |  |  | *Pseudemys peninsularis* | 1 |
|  |  |  |  | *Pyxis arachnoides* | 1 |
|  |  |  |  | *Pyxis planicauda* | 1 |
|  |  |  |  | *Rhinoclemmys areolata* | 1 |
|  |  |  |  | *Rhinoclemmys punctularia* | 1 |
|  |  |  |  | *Sacalia pseudocellata* | 1 |
|  |  |  |  | *Siebenrockiella crassicollis* | 1 |
|  |  |  |  | *Staurotypus salvinii* | 1 |
|  |  |  |  | *Staurotypus triporcatus* | 1 |
|  |  |  |  | *Sternotherus carinatus* | 1 |
|  |  |  |  | *Sternotherus odoratus* | 1 |
|  |  |  |  | *Terrapene carolina* | 1 |
|  |  |  |  | *Terrapene coahuila* | 1 |
|  |  |  |  | *Terrapene ornata* | 1 |
|  |  |  |  | *Testudo hermanni* | 1 |
|  |  |  |  | *Testudo kleinmanni* | 1 |
|  |  |  |  | *Testudo marginata* | 1 |
|  |  |  |  | *Trachemys dorbignyi* | 1 |
|  |  |  | African spurred tortoise | *Geochelone sulcate* | 1 |
|  |  |  | Alligator snapping turtle | *Macroclemys temminckii* | 1 |
|  |  |  | Asian forest tortoise | *Manouria emys* | 2 |
|  |  |  | Asian giant softshell turtle | *Pelochelys bibroni* | 2 |
|  |  |  | Asian leaf turtle | *Cyclemys dentata* | 2 |
|  |  |  | Asiatic soft shelled turtle | *Amyda cartilaginea* | 1 |
|  |  |  | Beale's eyed turtle | *Sacalia bealei* | 3 |
|  |  |  | Big headed turtle | *Platysternon megacephalum* | 4 |
|  |  |  | Black breasted leaf turtle | *Geoemyda spengleri* | 5 |
|  |  |  | Black pond turtle | *Geoclemys hamiltonii* | 2 |
|  |  |  | Black spine-necked swamp turtle | *Acanthochelys spixii* | 1 |
|  |  |  | Chinese box turtle | *Cuora flavomarginata* | 2 |
|  |  |  | Chinese broad headed pond turtle | *Chinemys megalocephala* | 1 |
|  |  |  | Chinese pond turtle | *Chinemys reevesii* | 4 |
|  |  |  | Chinese softshell turtle | *Pelodiscus sinensis* | 3 |
|  |  |  | Chinese softshell turtle | *Trionyx sinensis* | 1 |
|  |  |  | Chinese striped necked turtle | *Ocadia sinensis* | 3 |
|  |  |  | Chinese three striped box turtle | *Cuora trifasciata* | 3 |
|  |  |  | Crowned river turtle | *Hardella thurjii* | 2 |
|  |  |  | Elongated tortoise | *Indotestudo elongata* | 3 |
|  |  |  | Flap shelled turtle | *Lissemys punctata* | 1 |
|  |  |  | Florida soft-shelled turtle | *Apalone ferox* | 1 |
|  |  |  | Four eyed turtle | *Sacalia quadriocellata* | 3 |
|  |  |  | Hawksbill turtle | *Eretmochelys imbricata* | 1 |
|  |  |  | Horsfield’s tortoise | *Testudo horsfieldii* | 1 |
|  |  |  | Impressed tortoise | *Manouria impressa* | 2 |
|  |  |  | Indian black turtle | *Melanochelys trijuga* | 1 |
|  |  |  | Indian eyed turtle | *Morenia petersi* | 2 |
|  |  |  | Keeled box turtle | *Pyxidea mouhotii* | 2 |
|  |  |  | Loggerhead sea turtle | *Caretta caretta* | 1 |
|  |  |  | Matamata | *Chelus fimbriatus* | 1 |
|  |  |  | Mekong snail eating turtle | *Malayemys subtrijuga* | 2 |
|  |  |  | Northern red bellied turtle | *Pseudemys rubriventris* | 2 |
|  |  |  | Olive Ridley sea turtle | *Lepidochelys olivacea* | 1 |
|  |  |  | Painted turtle | *Chrysemys picta* | 2 |
|  |  |  | Pig nosed turtle | *Carettochelys insculpta* | 2 |
|  |  |  | Red eared slider | *Trachemys scripta elegans* | 2 |
|  |  |  | Ringed map turtle | *Graptemys oculifera* | 2 |
|  |  |  | River cooter | *Pseudemys concinna* | 2 |
|  |  |  | Smooth soft-shelled turtle | *Apalone mutica* | 1 |
|  |  |  | Southeast asian box turtle | *Cuora amboinensis* | 2 |
|  |  |  | Spiny soft-shelled turtle | *Apalone spinifera* | 1 |
|  |  |  | Spur thighed tortoise | *Testudo graeca* | 1 |
|  |  |  | Vietnamese box turtle | *Cuora galbinifrons* | 2 |
|  |  |  | Vietnamese box turtle | *Cuora galbinifrons* | 1 |
|  |  |  | Wattle necked softshell turtle | *Palea steindachneri* | 2 |
|  |  |  | Wattle necked softshell turtle | *Trionyx steindachneri* | 1 |
|  |  |  | Yellow blotched map turtle | *Graptemys flavimaculata* | 2 |
|  |  |  | Yellow pond turtle | *Mauremys mutica* | 3 |
|  |  |  | Yellow-spotted river turtle | *Podocnemis unifilis* | 1 |
|  |  |  | Yunnan box turtle | *Cuora yunnanensis* | 1 |
|  |  |  | Yunnan box turtle | *Cuora zhoui* | 1 |
| Europe | Reptilia | Squamata | Bell’s dabb lizard | *Uromastryx acanthinura* | 1 |
|  |  |  | Common chameleon | *Chamaeleo chamaeleon* | 2 |
|  |  |  | Desert monitor lizard | *Varanus griseus* | 1 |
|  |  |  | Egyptian cobra | *Naja haje* | 1 |
|  |  | Testudines | Spur thighed tortoise | *Testudo graeca* | 1 |
| Indian subcontinent | Reptilia | Testudines | Flap shelled turtle | *Lissemys punctata* | 1 |
|  |  |  | Indian black turtle | *Melanochelys trijuga* | 1 |
| Middle East | Aves | Accipitriformes | Griffon vulture | *Gyps fulvus* | 1 |
|  |  |  | Osprey | *Pandion haliaetus* | 1 |
|  |  | Anseriformes | Mallard | *Anas platyrhynchos* | 1 |
|  |  |  | Northern shoveler | *Anas clypeat* | 1 |
|  |  |  | Northern pintail | *Anas acuta* | 1 |
|  |  |  | Eurasian teal | *Anas crecca* | 1 |
|  |  | Charadriiformes | Eurasian thick knee | *Burhinus oedicnemus* | 1 |
|  |  | Columbiformes | Eurasian collared-dove | *Streptopelia decaocto* | 1 |
|  |  |  | Laughing dove | *Spilopelia senegalensis* | 1 |
|  |  | Falconiformes | Gyrfalcon | *Falco rusticolus* | 1 |
|  |  |  | Peregrine falcon | *Falco peregrinus* | 1 |
|  |  |  | Saker falcon | *Falco cherrug* | 1 |
|  |  | Galliformes | Arabian partridge | *Alectoris melanocephala* | 1 |
|  |  |  | Chukar | *Alectoris chukar* | 1 |
|  |  |  | Common quail | *Coturnix coturnix* | 1 |
|  |  |  | Sand partridge | *Ammoperdix heyi* | 1 |
|  |  |  | Common crane | *Grus grus* | 1 |
|  |  | Otidiformes | Houbara bustard | *Chlamydotis undulata* | 1 |
|  |  | Passeriformes | House sparrow | *Passer domesticus* | 1 |
|  |  |  | White eared bulbul | *Pycnonotus leucotis* | 1 |
|  |  | Psittaciformes | Blue and yellow macaw | *Ara ararauna* | 1 |
|  |  |  | Blue fronted amazon parrot | *Amazona aestiva* | 1 |
|  |  |  | Grey parrot | *Psittacus erithacus* | 1 |
|  |  |  | Rose ringed parakeet | *Psittacula krameri* | 1 |
|  |  |  | Yellow-crowned amazon | *Amazona ochrocephala* | 1 |
|  |  | Rheiformes | Greater rhea | *Rhea americana* | 1 |
|  | Mammalia | Artiodactyla | Nubian ibex | *Capra nubiana* | 1 |
|  |  | Carnivora | Arabian wolf | *Canis lupus* | 1 |
|  |  |  | Sand cat | *Felis margarita* | 1 |
|  |  | Hyracoidea | Rock hyrax | *Procavia capensis* | 1 |
|  |  | Lagomorpha | Brown hare | *Lepus capensis* | 1 |
|  | Reptilia | Squamata | Egyptian spiny tailed lizard | *Uromastyx aegyptia* | 1 |
|  |  | Testudines | Hawksbill turtle | *Eretmochelys imbricata* | 1 |
|  |  |  | Spur thighed tortoise | *Testudo graeca* | 1 |
|  |  |  | Western Caspian turtle | *Mauremys rivulata* | 1 |
| Oceania | Reptilia | Testudines | Pig nosed turtle | *Carettochelys insculpta* | 1 |
| South America | Amphibia | Anura | Marbled water frog | *Telmatobius marmoratus* | 1 |
|  | Aves | Passeriformes | Band tailed manakin | *Pipra fasciicauda* | 1 |
|  |  |  | Bearded bellbird | *Procnias averano* | 1 |
|  |  |  | Black faced tanager | *Schistochlamys melanopis* | 1 |
|  |  |  | Black throated saltator | *Saltatricula atricollis* | 1 |
|  |  |  | Blue black grassquit | *Volatinia jacarina* | 1 |
|  |  |  | Blue dacnis | *Dacnis cayana* | 1 |
|  |  |  | Brazilian tanager | *Ramphocelus bresilius* | 1 |
|  |  |  | Burnished buff tanager | *Tangara cayana* | 1 |
|  |  |  | Campo troupial | *Icterus jamacaii* | 1 |
|  |  |  | Chalk browed mockingbird | *Mumus saturninus* | 1 |
|  |  |  | Chestnut bellied seed finch | *Sporophila angolensis* | 1 |
|  |  |  | Chestnut capped blackbird | *Chrysomus ruficapillus* | 1 |
|  |  |  | Chopi blackbird | *Gnorimopsar chopi* | 1 |
|  |  |  | Cinnamon tanager | *Schistochlamys ruficapillus* | 1 |
|  |  |  | Common waxbill | *Estrilda astrild* | 1 |
|  |  |  | Copper seedeater | *Sporophila bouvreuil* | 1 |
|  |  |  | Creamy bellied thrush | *Turdus amaurochalinus* | 1 |
|  |  |  | Crested oropendola | *Psarocolius decumanus* | 1 |
|  |  |  | Double collared seedeater | *Sporophila caerulescens* | 1 |
|  |  |  | Epaulet oriole | *Icterus cayanensis* | 1 |
|  |  |  | Green winged saltator | *Saltator similis* | 1 |
|  |  |  | Grey pileated finch | *Lanio pileatus* | 1 |
|  |  |  | Guira tanager | *Hemithraupis guira* | 1 |
|  |  |  | Hooded siskin | *Sporagra magellanica* | 1 |
|  |  |  | Lined seedeater | *Sporophila lineola* | 1 |
|  |  |  | Pale breasted thrush | *Turdus leucomelas* | 1 |
|  |  |  | Palm tanager | *Tangara palmarum* | 1 |
|  |  | Passeriformes | Pectoral sparrow | *Arremon taciturnus* | 1 |
|  |  |  | Plumbeous seedeater | *Sporophila plumbea* | 1 |
|  |  |  | Purple throated euphonia | *Euphonia chlorotica* | 1 |
|  |  |  | Red cowled cardinal | *Paroaria dominicana* | 1 |
|  |  |  | Red necked tanager | *Tangara cyanocephala* | 1 |
|  |  |  | Red pileated finch | *Lanio cucullatus* | 1 |
|  |  |  | Rufous bellied thrush | *Turfus rufiventris* | 1 |
|  |  |  | Saffron finch | *Sicalis flaveola* | 2 |
|  |  |  | Scarlet throated tanager | *Compsothraupis loricata* | 1 |
|  |  |  | Shiny cowbird | *Molothrus bonariensis* | 1 |
|  |  |  | Solitary cacique | *Procacicus solitarius* | 1 |
|  |  |  | Tangara sayaca | *Thraupis sayaca* | 1 |
|  |  |  | Tropical mockingbird | *Mimus gilvus* | 1 |
|  |  |  | Ultramarine grosbeak | *Cyanoloxia brissonii* | 1 |
|  |  |  | Violaceous euphonia | *Euphonia violacea* | 1 |
|  |  |  | White browed meadowlark | *Sturnella superciliaris* | 1 |
|  |  |  | White naped jay | *Cyanocorax cyanopogon* | 1 |
|  |  |  | White throated seedeater | *Sporophila alborgularis* | 1 |
|  |  |  | Wild canary | *Serinus canaria* | 1 |
|  |  |  | Yellow bellied seedeater | *Sporophila nigricollis* | 1 |
|  |  |  | Yellow faced siskin | *Sporagra yarrellii* | 1 |
|  |  | Piciformes | Gould's toucanet | *Selenidera gouldii* | 1 |
|  |  | Psittaciformes | Blue fronted amazon parrot | *Amazona aestiva* | 2 |
|  |  |  | Blue headed parrot | *Pionus menstruus* | 1 |
|  |  |  | Blue throated parakeet | *Pyrrhura cruentata* | 1 |
|  |  |  | Blue winged macaw | *Primolius maracana* | 1 |
|  |  |  | Blue winged parrotlet | *Forpus xanthopterygius* | 1 |
|  |  |  | Caatinga parakeet | *Aratinga cactorum* | 1 |
|  |  |  | Crimson bellied parakeet | *Pyrrhura perlata* | 1 |
|  |  |  | Grey breasted parakeet | *Pyrrhura griseipectus* | 1 |
|  |  |  | Jandaya parakeet | *Aratinga jandaya* | 1 |
|  |  |  | Red and green macaw | *Ara chloropterus* | 1 |
|  |  |  | Rose fronted parakeet | *Pyrrhura roseifrons* | 1 |
|  |  |  | Rose ringed parakeet | *Psittacula krameri* | 1 |
|  |  |  | Yellow chevroned parakeet | *Brotogeris chiriri* | 1 |
|  |  |  | Yellow rumped cacique | *Cacicus cela* | 1 |
|  | Mammalia | Cingulata | Hairy armadillo | *Chaetophractus nationi* | 1 |
|  |  | Didelphimorphia | Common opossum | *Didelphis marsupialis* | 1 |
|  | Reptilia | Squamata | Boa constrictor | *Boa constrictor* | 1 |
|  |  | Testudines | Matamata | *Chelus fimbriatus* | 1 |
|  |  |  | Red footed tortoise | *Chelonoidis carbonaria* | 1 |
|  |  |  | Yellow footed tortoise | *Chelonoidis denticulata* | 2 |
| Southeast Asia | Amphibia | Anura | American bullfrog | *Rana catesbeiana* | 1 |
|  |  |  | Asian grass frog | *Fejervarya limnocharis* | 1 |
|  |  |  | Crab eating frog | *Fejervarya cancrivora* | 1 |
|  |  |  | Dark spotted frog | *Rana nigromaculata* | 1 |
|  |  |  | East Asian bullfrog | *Hoplobatrachus rugulosus* | 1 |
|  |  |  | Fanged river frog | *Limnonectes macrodon* | 1 |
|  | Aves | Anseriformes | Mallard | *Anas platyrhynchos* | 1 |
|  |  | Bucerotiformes | Rufous necked hornbill | *Aceros nipalensis* | 1 |
|  |  | Columbiformes | Grey capped emerald dove | *Chalcophaps indica* | 1 |
|  |  |  | Spotted dove | *Streptopelia chinensis* | 1 |
|  |  |  | Zebra dove | *Geopelia striata* | 1 |
|  |  | Cuculiformes | Greater coucal | *Centropus sinensis* | 1 |
|  |  | Galliformes | Chinese francolin | *Francolinus pintadeanus* | 1 |
|  |  |  | Common pheasant | *Phasianus colchicus* | 1 |
|  |  |  | Grey peacock pheasant | *Polyplectron bicalcaratum* | 1 |
|  |  | Gruiformes | White breasted waterhen | *Amaurornis phoenicurus* | 2 |
|  |  | Passeriformes | Asian fairy bluebird | *Irena puella* | 1 |
|  |  |  | Asian glossy starling | *Aplonis panayensis strigata* | 1 |
|  |  |  | Asian pied starling | *Gracupica contra* | 1 |
|  |  |  | Bar winged prinia | *Prinia familiaris* | 1 |
|  |  |  | Baya weaver | *Ploceus philippinus* | 1 |
|  |  |  | Black capped white eye | *Zosterops atricapilla* | 1 |
|  |  |  | Black collared starling | *Sturnus nigricollis* | 1 |
|  |  |  | Black headed bulbul | *Pycnonotus atriceps atriceps* | 1 |
|  |  |  | Blue masked leafbird | *Chloropsis venusta* | 1 |
|  |  |  | Blue winged leafbird | *Chloropsis moluccensis* | 1 |
|  |  |  | Brown throated sunbird | *Anthreptes malacensis* | 1 |
|  |  |  | Chestnut capped laughing thrush | *Garrulax mitratus* | 1 |
|  |  |  | Chestnut munia | *Lonchura atricapilla* | 1 |
|  |  |  | Cinereous tit | *Parus cinereus* | 1 |
|  |  |  | Common hill mynah | *Gracula religiosa* | 1 |
|  |  |  | Common iora | *Aegithina tiphia scapularis* | 1 |
|  |  |  | Common mynah | *Acridotheres tristis* | 1 |
|  |  |  | Daurian starling | *Agropsar sturninus* | 1 |
|  |  |  | Finch billed mynah | *Scissirostrum dubium* | 1 |
|  |  |  | Greater green leafbird | *Chloropsis sonnerati sonnerati* | 1 |
|  |  |  | Hwamei | *Garrulax canorus* | 1 |
|  |  |  | Java sparrow | *Padda oryzivora* | 1 |
|  |  |  | Javan mynah | *Acridotheres javanicus* | 1 |
|  |  |  | Lesser green leafbird | *Chloropsis cyanopogon* | 1 |
|  |  |  | Long tailed shrike | *Lanius schach bentet* | 1 |
|  |  |  | Long tailed sibia | *Heterophasia picaoides* | 1 |
|  |  |  | Ochraceous bulbul | *Alophoixus ochraceus sumatranus* | 1 |
|  |  |  | Olive backed sunbird | *Cinnyris jugularis* | 1 |
|  |  |  | Olive winged bulbul | *Pycnonotus plumosus* | 1 |
|  |  |  | Orange bellied flowerpecker | *Dicaeum trigonostigma flaviclunis* | 1 |
|  |  |  | Oriental magpie robin | *Copsychus saularis muticus pluto* | 1 |
|  |  |  | Oriental white eye | *Zosterops palpebrosus* | 1 |
|  |  |  | Ruby throated bulbul | *Pycnonotus dispar* | 1 |
|  |  |  | Sangkar white eye | *Zosterops melanurus buxtoni* | 1 |
|  |  |  | Scaly breasted munia | *Lonchura punctulata* | 1 |
|  |  |  | Scarlet minivet | *Pericrocotus flammeus* | 1 |
|  |  |  | Sooty headed bulbul | *Pycnonotus aurigaster* | 1 |
|  |  |  | Streaked bulbul | *Ixos malaccensis* | 1 |
|  |  |  | Sumatran bulbul | *Ixos sumatranus* | 1 |
|  |  |  | White headed munia | *Lonchura maja* | 1 |
|  |  |  | White rumped shama | *Kittacincla malabarica tricolor* | 1 |
|  |  |  | Yellow vented bulbul | *Pycnonotus goiavier analis* | 1 |
|  |  | Piciformes | Common flameback | *Dinopium javanense* | 1 |
|  |  | Psittaciformes | Blue crowned hanging parrot | *Loriculus galgulus* | 1 |
|  |  |  | Coconut lorikeet | *Trichoglossus haematodus* | 1 |
|  |  |  | Red breasted parakeet | *Psittacula alexamdri* | 1 |
|  |  |  | Red lory | *Eos bornea* | 1 |
|  |  | Strigiformes | Eurasian eagle owl | *Bubo bubo* | 1 |
|  | Mammalia | Artiodactyla | Hog deer | *Axis porcinus* | 1 |
|  |  | Carnivora | Crab eating mongoose | *Herpestes urva* | 1 |
|  |  |  | Jungle cat | *Felis chaus* | 1 |
|  |  |  | Large Indian civet | *Viverra zibetha* | 1 |
|  |  |  | Leopard cat | *Felis bengalensis* | 1 |
|  |  |  | Lesser Indian civet | *Viverricula indica* | 1 |
|  |  |  | Masked palm civet | *Paguma larvata* | 1 |
|  |  | Pholidota | Chinese pangolin | *Manis pentadactyla* | 2 |
|  |  |  | Indian pangolin | *Manis crassicaudata* | 1 |
|  |  |  | Sunda pangolin | *Manis javanica* | 1 |
|  |  | Primates | Assam macaque | *Macaca assamensis* | 2 |
|  |  |  | Francois' leaf monkey | *Trachypithecus francoisi* | 1 |
|  |  |  | Long tailed macaque | *Macaca fascicularis* | 1 |
|  |  |  | Pig tailed macaque | *Macaca nemestrina* | 1 |
|  |  |  | Rhesus macaque | *Macaca mulatta* | 1 |
|  |  |  | Sunda slow loris | *Nycticebus coucang* | 3 |
|  |  |  | Stump tailed macaque | *Macaca arctoides* | 1 |
|  |  |  | Tonkin snub nosed monkey | *Pygathrix avunculus* | 1 |
|  |  | Rodentia | Large bamboo rat | *Rhizomys sumatrensis* | 1 |
|  |  |  | Giant flying squirrel | *Petaurista philippensis* | 1 |
|  |  |  | Hoary bamboo rat | *Rhizomys pruinosus* | 1 |
|  |  |  | Laotian rock rat | *Laonastes aenigmamus* | 1 |
|  |  |  | Pallas's squirrel | *Callosciurus erythraeus* | 1 |
|  |  |  | Asiatic brush-tailed porcupine | *Atherurus macrourus* | 1 |
|  | Reptilia | Squamata | Asian cobra | *Naja naja* | 1 |
|  |  |  | Asian rock python | *Python molurus* | 1 |
|  |  |  | Banded krait | *Bungarus fasciatus* | 1 |
|  |  |  | Chinese rat snake | *Pytas korros* | 2 |
|  |  |  | Chinese water dragon | *Physignathus cocincinus* | 3 |
|  |  |  | Clouded monitor | *Varanus nebulosus* | 2 |
|  |  |  | Common sun skink | *Eutropis multifasciata* | 1 |
|  |  |  | Green snake | *Opheodrys major* | 1 |
|  |  |  | Indonesian bronzeback snake | *Dendrelaphis pictus* | 1 |
|  |  |  | King cobra | *Ophiophagus hannah* | 1 |
|  |  |  | Oriental garden lizard | *Calotes versicolor* | 2 |
|  |  |  | Oriental rat snake | *Ptyas mucosus* | 1 |
|  |  |  | Pit viper | *Deinagkistrodon acutus* | 1 |
|  |  |  | Rainbow watersnake | *Enhydris enhydris* | 1 |
|  |  |  | Radiated ratsnake | *Elaphe radiata* | 1 |
|  |  |  | Slender sea snake | *Microcephalophis gracilis* | 1 |
|  |  |  | Striped watersnake | *Enhydris jagorii* | 1 |
|  |  |  | Thin ground snake | *Elaphe taeniurus* | 1 |
|  |  |  | Tokay gecko | *Gecko gecko* | 1 |
|  |  |  | Water monitor | *Varanus salvator* | 2 |
|  |  | Testudines | Alligator snapping turtle | *Macrochelys temminckii* | 1 |
|  |  |  | Asiatic soft shelled turtle | *Amyda cartilaginea* | 1 |
|  |  |  | Big headed turtle | *Platysternon megacephalum* | 2 |
|  |  |  | Black breasted leaf turtle | *Geoemyda spengleri* | 1 |
|  |  |  | Black marsh turtle | *Siebenrockiella crassicollis* | 1 |
|  |  |  | Chinese pond turtle | *Chinemys reevesii* | 1 |
|  |  |  | Chinese softshell turtle | *Pelodiscus sinensis* | 2 |
|  |  |  | Chinese softshell turtle | *Trionyx sinensis* | 1 |
|  |  |  | Chinese striped necked turtle | *Ocadia sinensis* | 1 |
|  |  |  | Chinese three striped box turtle | *Cuora trifasciata* | 1 |
|  |  |  | Elongated tortoise | *Indotestudo elongata* | 1 |
|  |  |  | Impressed tortoise | *Manouria impressa* | 2 |
|  |  |  | Loggerhead sea turtle | *Caretta caretta* | 1 |
|  |  |  | Malayan snail eating turtle | *Malayemys macrocephala* | 1 |
|  |  |  | Mekong snail eating turtle | *Malayemys subtrijuga* | 2 |
|  |  |  | Oldham's leaf turtle | *Cyclemys oldhamii* | 1 |
|  |  |  | Olive Ridley sea turtle | *Lepidochelys olivacea* | 1 |
|  |  |  | Ornate softshell turtle | *Amyda ornate* | 1 |
|  |  |  | Red eared slider | *Trachemys scripta elegans* | 1 |
|  |  |  | Southeast Asian box turtle | *Cuora amboinensis* | 2 |
|  |  |  | Wattle necked softshell turtle | *Trionyx steindachneri* | 1 |

Table S2 List of live, vertebrate, terrestrial wildlife that were not differentiated to species, reported in markets in a scoping review of research associated with live, terrestrial, vertebrate wildlife sold for any purpose at markets likely to sell food.

| Region | Live wildlife | Count |
| --- | --- | --- |
| Africa | Bird egg | 1 |
|  | Cane rat (Thryonomys) | 1 |
|  | Crocodile (Crocodylus) | 1 |
|  | Monitor lizard (Varanus) | 1 |
|  | Pangolin (Phataginus) | 2 |
|  | Reptile egg | 1 |
|  | Tortoise (Kinixys) | 2 |
| China | Duck (Anas) | 1 |
|  | Lark (Allaudidae) | 1 |
|  | Partridge (Arborophila) | 1 |
|  | Plain long nosed squirrel (Dremomys) | 1 |
|  | Accipiteridae | 1 |
|  | Axolotl | 1 |
|  | Flying squirrel (Petaurista) | 1 |
|  | Grosbeak (Pinicola) | 1 |
|  | Knobby newt (Tylotoriton) | 1 |
|  | Leiothrix | 1 |
|  | Macaque | 1 |
|  | Monkey | 1 |
|  | Mynah | 1 |
|  | Old world oriole (Oriolus) | 1 |
|  | Owl (Tytonidae) | 1 |
|  | Pangolin (Manis) | 1 |
|  | Parrots | 1 |
|  | Pelomedusidae | 1 |
|  | Porcupine (Hystricoidea) | 1 |
|  | Snake | 2 |
|  | Sparrow | 1 |
|  | Squirrel (Tamiops) | 2 |
|  | Testudinidae | 2 |
|  | Thrush | 1 |
|  | Tit (Parus) | 2 |
|  | Toad (Bufo) | 1 |
| Southeast Asia | Accipiteridae | 1 |
|  | Bear (Ursidae) | 1 |
|  | Cobra (Naja) | 1 |
|  | Lark (Alaudidae) | 1 |
|  | Leaf warbler (Phylloscopus) | 1 |
|  | Munia (Lonchura) | 1 |
|  | Owl (Tytonidae) | 1 |
|  | Pangolin (Manis) | 2 |
|  | Testudinidae | 1 |
